# Blood Proteomic Biomarkers indicate reduced system and tissue level inflammation responses to The GOTO Lifestyle Intervention in Older Adults

**DOI:** 10.1101/2025.10.08.25337394

**Authors:** F.A. Bogaards, T. Gehrmann, M. Beekman, N. Lakenberg, E. Suchiman, R.-U. Müller, J. Deelen, J.-W. Lackmann, S. Müller, C.P.G.M. de Groot, M.J.T. Reinders, P. Antczak, P.E. Slagboom

## Abstract

The effects of lifestyle interventions on the blood metabolome and transcriptome are well studied, but the interplay between tissues is not well understood. In a multi-omics multi-tissue design we study how the serum proteome reflects health improvements in age-related deficits such as body composition, fat and glucose metabolism and low-grade inflammation in older adults undergoing a 13-week lifestyle intervention (GOTO trial). Significant intervention effects on the fasting blood proteome were observed in both sexes, with downregulated immune-related pathways, including the complement system, inflammation, cholesterol transport and coagulation. These findings associated with improved body composition and immune-metabolic health markers. In addition, we found that the intervention-induced changes in the transcriptome of muscle and subcutaneous adipose tissue (SAT) biopsies were correlated with changes of the corresponding circulating proteins. The most notable correlations were observed for proteins involved in inflammatory processes and corresponding gene expression in the SAT. Our findings emphasize the broad beneficial effects of moderate lifestyle interventions in older adults and how circulating proteins such as FN1, LGALS3BP and PRG4 reflect the immune-metabolic health benefits.

## Introduction

Over the last 200 years, the life expectancy has steadily increased in most developed nations^1^. Concomitantly, the proportion of the population aged over 60 increased and is expected to rise from 12% in 2015 to 22% in 2050 ^2^. This appears as a clear success at first sight, which, however, leads to a significant increase in disease burden and healthcare costs ^1^. Increasing actual healthspan would be key to solving this problem. One of the main risk factors for the age-related health decline is systemic chronic low-grade inflammation ^3^, contributing to a plethora of age-related diseases including cardiovascular, renal, metabolic, neurodegenerative and immune system diseases ^3^. To maintain vitality and independence in older ages, it is crucial to attenuate the age-related health decline and tackle systemic chronic inflammation. Indeed, lifestyle interventions, through a change in diet, physical activity or a combination of the two, have shown to improve metabolic and immune-related health parameters ^4–6^. The responses to interventions can be heterogeneous, especially in older adults, highlighting the necessity for more in-depth research to enable personalized approaches is required ^4,7–12^.

The health gain in lifestyle interventions can be tracked by performing comprehensive measurements at both baseline and after the intervention. Typical measurements included are anthropometrics (i.e., weight, height, waist circumference), traditional blood-based markers of health (i.e., fasting insulin level, glucose level, fasting C-reactive protein), and omics-based analyses (i.e., DNA methylation, transcriptomics, proteomics and metabolomics). Several studies have reported on lifestyle responses of the blood DNA methylome ^13–15^, transcriptome ^9,16–18^, and metabolome ^4,19–22^. Current literature does indicate that the serum metabolome strongly responds to lifestyle changes ^4,11,19–22^, contrary to the whole blood transcriptome, which only seems to respond to intense lifestyle interventions ^10,16–18^. This indicates that transcriptome of whole blood is likely not the ideal molecular measurement to monitor the response to a mild lifestyle intervention.

In recent years, it has become more evident that only part of the proteins measured in blood are produced in blood, generally as part of the immune response ^23^, the rest being representative of other tissues and either actively or inactively secreted into the blood stream^24^. Moreover, a recent study has shown that plasma proteins enriched in specific organs can be indicative of organ ageing ^25^. These developments highlight the potential for the circulating proteome in biomarker development. However, whether this data source can also represent tissue-specific changes induced by a mild lifestyle intervention or whether baseline values of these proteins can be indicative of a stronger health benefit has not been studied in depth. In the current study, we therefore investigated the effects of a 13-week combined lifestyle intervention on the fasting blood proteome of older adults. We related the changes of the intervention responding proteins (IRPs) to changes in a range of health markers reflecting blood pressure, body composition, glucose and fat metabolism, as well as inflammation. In addition, we explored whether the intervention-induced changes of the fasting serum proteome only reflected previously observed changes in the postprandial transcriptome of blood, or also in the transcriptomes of subcutaneous adipose tissue (SAT) and muscle tissue ^16^.

## Results

### GOTO lifestyle intervention trial

The Growing Old TOgether (GOTO) trial, was a 13-week combined lifestyle intervention with 164 participants, of whom 163 completed the trial. One participant withdrew because of a knee surgery ^4,16^. Of the 163 participants, 153 were considered compliant according to the compliance thresholds described in Bogaards et al ^16^. Of these 153 compliant participants, the fasting blood proteome was measured in 146 (70 male, 76 female). The intervention consisted of 12.5% reduced caloric intake and 12.5% increased physical activity. The primary outcome of the trial was a significant reduction in fasting insulin^4^ and, in addition, several secondary outcomes including blood parameters, anthropometrics, body composition and blood pressure were measured^16^.

### The GOTO trial improved the metabolic health status of participants

For the current study, we focused on the immune-metabolic health of the GOTO participants, at baseline and after the intervention by looking at some of the health markers for which we previously showed that they are changed by the lifestyle intervention ^4^, namely *I)* blood pressure: systolic blood pressure (SBP) and diastolic blood pressure (DBP), *II)* body composition: body mass index (BMI), waist circumference (WC), total body fat % and trunk fat%, *III)* fasted state serum markers of fat metabolism: low-density lipoprotein (LDL) cholesterol, high-density lipoprotein (HDL) cholesterol, HDL cholesterol size and serum triglycerides (TG), *IV)* fasted state serum markers of glucose metabolism: insulin (the primary outcome of the GOTO trial) and glucose, *V)* fasted state serum markers of inflammation: C-reactive protein (CRP) and alpha-1-acid glycoprotein (GlycA), a novel low grade inflammation marker ^26^. These health markers were studied in male and female participants separately.

At baseline, the participants had a mean age of 64.1 and 62.0 yrs, and an average BMI of 26.6 and 27.0 kg/m^2^ for males and females, respectively (Table 1). Most of the baseline health marker values did not differ significantly between the male and female participants, including fasting insulin. Some measurements, however, did show significant sex differences: total body fat %, trunk fat % and both HDL cholesterol concentration and size were higher in female participants, whereas WC was higher in the male participants.

**Table 1:**
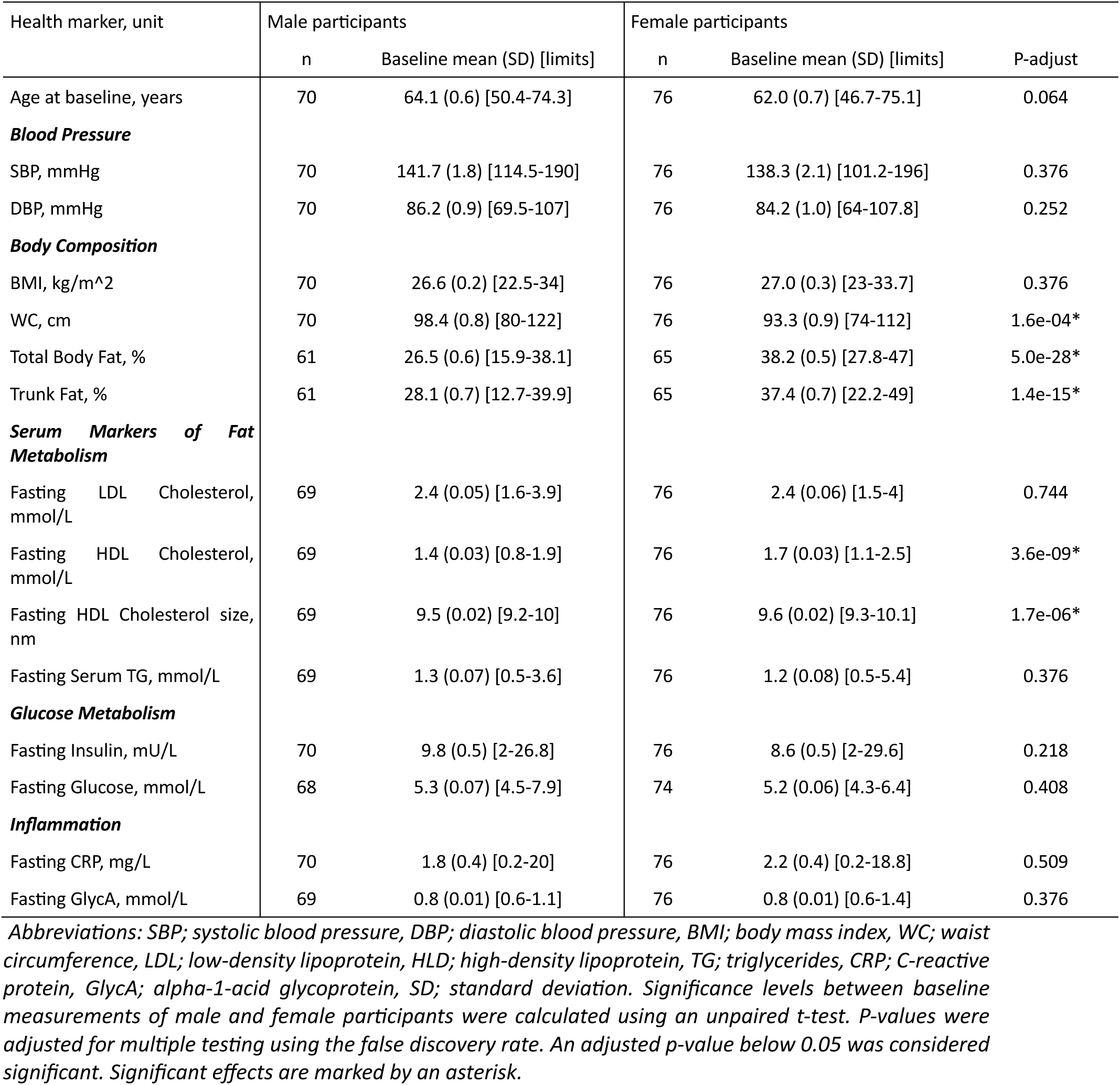
Baseline immune-metabolic health markers of the GOTO trial participants with fasting blood proteome measurements, stratified by sex.

| Health marker, unit | Male participants |  | Female participants |  |  |
| --- | --- | --- | --- | --- | --- |
|  | n | Baseline mean (SD) [limits] | n | Baseline mean (SD) [limits] | P-adjust |
| Age at baseline, years | 70 | 64.1 (0.6) [50.4-74.3] | 76 | 62.0 (0.7) [46.7-75.1] | 0.064 |
| **Blood Pressure** |  |  |  |  |  |
| SBP, mmHg | 70 | 141.7 (1.8) [114.5-190] | 76 | 138.3 (2.1) [101.2-196] | 0.376 |
| DBP, mmHg | 70 | 86.2 (0.9) [69.5-107] | 76 | 84.2 (1.0) [64-107.8] | 0.252 |
| **Body Composition** |  |  |  |  |  |
| BMI, kg/m<sup>2</sup> | 70 | 26.6 (0.2) [22.5-34] | 76 | 27.0 (0.3) [23-33.7] | 0.376 |
| WC, cm | 70 | 98.4 (0.8) [80-122] | 76 | 93.3 (0.9) [74-112] | 1.6e-04* |
| Total Body Fat, % | 61 | 26.5 (0.6) [15.9-38.1] | 65 | 38.2 (0.5) [27.8-47] | 5.0e-28* |
| Trunk Fat, % | 61 | 28.1 (0.7) [12.7-39.9] | 65 | 37.4 (0.7) [22.2-49] | 1.4e-15* |
| **Serum Markers of Fat Metabolism** |  |  |  |  |  |
| Fasting LDL Cholesterol, mmol/L | 69 | 2.4 (0.05) [1.6-3.9] | 76 | 2.4 (0.06) [1.5-4] | 0.744 |
| Fasting HDL Cholesterol, mmol/L | 69 | 1.4 (0.03) [0.8-1.9] | 76 | 1.7 (0.03) [1.1-2.5] | 3.6e-09* |
| Fasting HDL Cholesterol size, nm | 69 | 9.5 (0.02) [9.2-10] | 76 | 9.6 (0.02) [9.3-10.1] | 1.7e-06* |
| Fasting Serum TG, mmol/L | 69 | 1.3 (0.07) [0.5-3.6] | 76 | 1.2 (0.08) [0.5-5.4] | 0.376 |
| **Glucose Metabolism** |  |  |  |  |  |
| Fasting Insulin, mU/L | 70 | 9.8 (0.5) [2-26.8] | 76 | 8.6 (0.5) [2-29.6] | 0.218 |
| Fasting Glucose, mmol/L | 68 | 5.3 (0.07) [4.5-7.9] | 74 | 5.2 (0.06) [4.3-6.4] | 0.408 |
| **Inflammation** |  |  |  |  |  |
| Fasting CRP, mg/L | 70 | 1.8 (0.4) [0.2-20] | 76 | 2.2 (0.4) [0.2-18.8] | 0.509 |
| Fasting GlycA, mmol/L | 69 | 0.8 (0.01) [0.6-1.1] | 76 | 0.8 (0.01) [0.6-1.4] | 0.376 |

Abbreviations: SBP; systolic blood pressure, DBP; diastolic blood pressure, BMI; body mass index, WC; waist circumference, LDL; low-density lipoprotein, HDL; high-density lipoprotein, TG; triglycerides, CRP; C-reactive protein, GlycA; alpha-1-acid glycoprotein, SD; standard deviation. Significance levels between baseline measurements of male and female participants were calculated using an unpaired t-test. P-values were adjusted for multiple testing using the false discovery rate. An adjusted p-value below 0.05 was considered significant. Significant effects are marked by an asterisk.

When focusing on only the people without lipid lowering medication or antihypertension medication at baseline, no significant differences were present between male and female participants for the blood pressure and fat metabolism markers (Supplementary Table 1).

The GOTO intervention had a positive effect on the overall immune-metabolic health of both sexes as reflected by significant changes in body composition, serum markers of fat metabolism and inflammation (Table 2), which is in line with what was shown previously using both sex-combined and stratified data ^4^. Of the two blood pressure measurements, SBP reduced significantly in male participants. In both sexes, all four body composition measurements reduced significantly. Of the fat metabolism measurements, LDL cholesterol reduced significantly in both sexes, whereas fasting HDL cholesterol size significantly increased both in males and females. Some significant effects were only present in male participants: a significant increase in fasting HDL cholesterol concentration and a significant decrease in serum TG and glucose. Notably, the effect of fasting HDL concentration in the female participants was trending negatively, in opposite direction to the effect in male participants. Fasting GlycA, one of the two inflammatory related health markers, was significantly reduced in both sexes. Fasting CRP, on the other hand, showed a non-significant decrease in both sexes. Similarly, the two blood pressure and the two glucose metabolism health markers all decreased, but not significantly, in both sexes. (Table 2). Overall, the intervention improved the immune-metabolic health status in both sexes, with slightly stronger effects in the male participants.

**Table 2:** Effects of the GOTO intervention on immune-metabolic health markers in participants with fasting blood proteome measurements.

| Health marker, unit | Male participants |  |  | Female participants |  |  |
| --- | --- | --- | --- | --- | --- | --- |
|  | n | Mean (SD) | p-adjust | n | Mean (SD) | p-adjust |
| **Blood Pressure** |  |  |  |  |  |  |
| SBP, mmHg | 70 | -3.5 (20.9) | 0.03* | 76 | -2.9 (23.2) | 0.058 |
| DBP, mmHg | 70 | -0.7 (10.4) | 0.425 | 76 | -1.3 (11.5) | 0.123 |
| **Body Composition** |  |  |  |  |  |  |
| BMI, kg/m<sup>2</sup> | 70 | -1.1 (2.7) | 1.7e-16* | 76 | -1.2 (3.3) | 1.7e-19* |
| WC, cm | 70 | -4.4 (10.1) | 1.1e-08* | 76 | -4.5 (10.1) | 5.6e-09* |
| Total Body Fat, % | 58 | -1.7 (5.7) | 1.1e-09* | 60 | -1.1 (5.3) | 7.5e-09* |
| Trunk Fat, % | 58 | -2.6 (7.3) | 1.5e-11* | 60 | -1.7 (6.7) | 2.4e-10* |
| **Serum Markers of Fat Metabolism** |  |  |  |  |  |  |
| Fasting LDL Cholesterol, mmol/L | 69 | -0.2 (0.6) | 6.1e-04* | 76 | -0.2 (0.5) | 7.2e-06* |
| Fasting HDL Cholesterol, mmol/L | 69 | 0.04 (0.3) | 0.024* | 76 | -0.04 (0.3) | 0.057 |
| Fasting HDL Cholesterol size, nm | 69 | 0.06 (0.2) | 3.6e-06* | 76 | 0.03 (0.2) | 0.004* |
| Fasting Serum TG, mmol/L | 69 | -0.2 (0.7) | 6.3e-04* | 76 | -0.1 (0.8) | 0.120 |
| **Glucose Metabolism** |  |  |  |  |  |  |
| Fasting Insulin, mU/L | 70 | -0.4 (6.1) | 0.327 | 76 | -0.3 (5.7) | 0.402 |
| Fasting Glucose, mmol/L | 64 | -0.1 (0.7) | 0.011* | 66 | -0.07 (0.6) | 0.171 |
| **Inflammation** |  |  |  |  |  |  |
| Fasting CRP, mg/L | 70 | -0.4 (3.6) | 0.327 | 76 | -0.5 (3.1) | 0.149 |
| Fasting GlycA, mmol/L | 69 | -0.03 (0.1) | 0.004* | 76 | -0.02 (0.1) | 0.035* |

Abbreviations: SBP; systolic blood pressure, DBP; diastolic blood pressure, BMI; body mass index, WC; waist circumference, LDL; low-density lipoprotein, HDL; high-density lipoprotein, TG; triglycerides, CRP; C-reactive protein, GlycA; alpha-1-acid glycoprotein, SD; standard deviation. SD; standard deviation. Significance levels were calculated using a two-sided linear mixed model. P-values were adjusted for multiple testing using the false discovery rate. An adjusted p-value below 0.05 was considered significant. Significant effects are marked by an asterisk.

The effects on blood pressure were more pronounced when the participants who were on antihypertensive medication were removed. The effects on fat metabolism markers were similar when the participants on lipid lowering medication were removed from the analysis. This resulted in significant reductions of SBP, DBP (female only), LDL cholesterol concentration, HDL cholesterol concentration (female only), serum triglycerides (male only) and an increase in HDL cholesterol size (Supplementary Table 2). It is interesting to note that while the change in HDL cholesterol was significant in the entire set of male participants, it was not in the subset of participants not receiving lipid-lowering medication. In female participants, the intervention appeared to have a negative effect on the concentration of HDL cholesterol, with an effect that approached significance in the total set of female participants (FDR = 0.057). When only selecting the participants without lipid lowering medication, this effect was significant.

Even though slight differences were present in the group without the participants taking antihypertensive and/or lipid lowering medication, we were interested in the effect of the intervention across all individuals regardless of medication usage.

### The GOTO intervention had minimal effects on cell type composition

To investigate whether the intervention changed the cell type composition, which in turn can influence the protein levels, we investigated the effect of the intervention on the cell type percentages of Neutrophiles, Basophiles, Eosinophiles, Lymphocytes, Monocytes and Leukocytes. None of these cell types significantly changed after multiple testing correction (Supplementary Table 3). However, at the nominal p-value threshold, the percentage of leukocytes was reduced in the male participants, we subsequently adjusted our proteome-based analyses for cell type composition.

### Sex-dependent proteomic response in the fasting blood proteome

We characterized 338 proteins in fasting blood (Table 3). To understand how the fasting blood proteome changes across the GOTO intervention, we performed a sex-stratified paired analysis for each participant. The overall pattern of the effect of the intervention was similar in males and females, with roughly 80% of the significantly intervention responding proteins (IRPs) downregulated (Figure 1, Table 3). However, we observed some differences in the effect sizes and significance levels between the two sexes.

**Figure 1:**
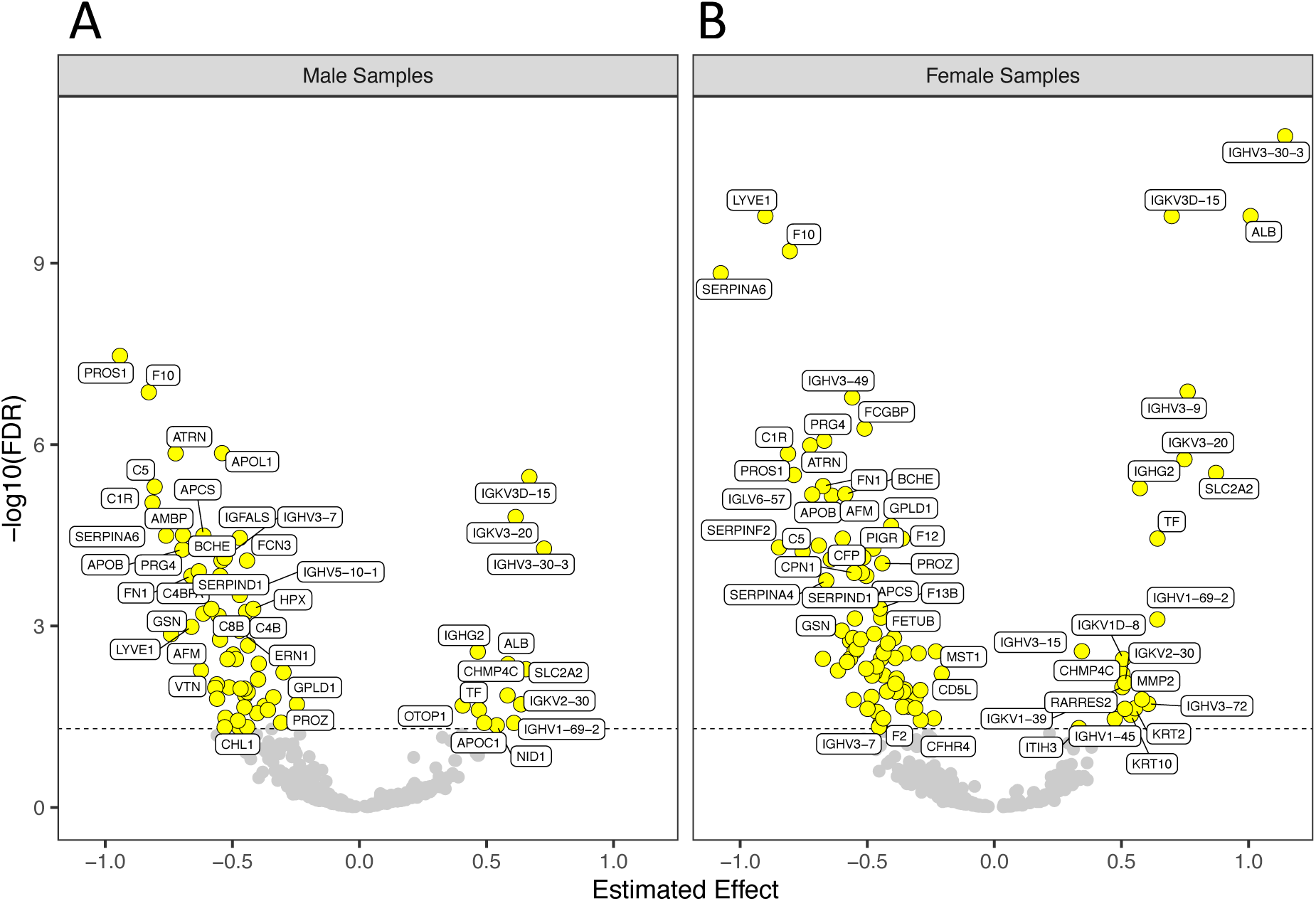
The intervention reduced protein levels involved in the innate immune system. Intervention effect on fasting blood proteome levels in males (A) and females (B). The effect of the intervention was studied using two-sided linear mixed model analysis. The significance levels were adjusted for multiple testing using the false discovery rate per sex (FDR). The estimated effect reflects the scaled intervention effect and is plotted on the x-axis. Proteins with an FDR adjusted p-value < 0.05 were considered significantly changed. Each point represents a protein. Yellow points represent the significantly changed proteins. Grey points represent the proteins with an FDR adjusted p-value ≥ 0.05.

**Table 3:**
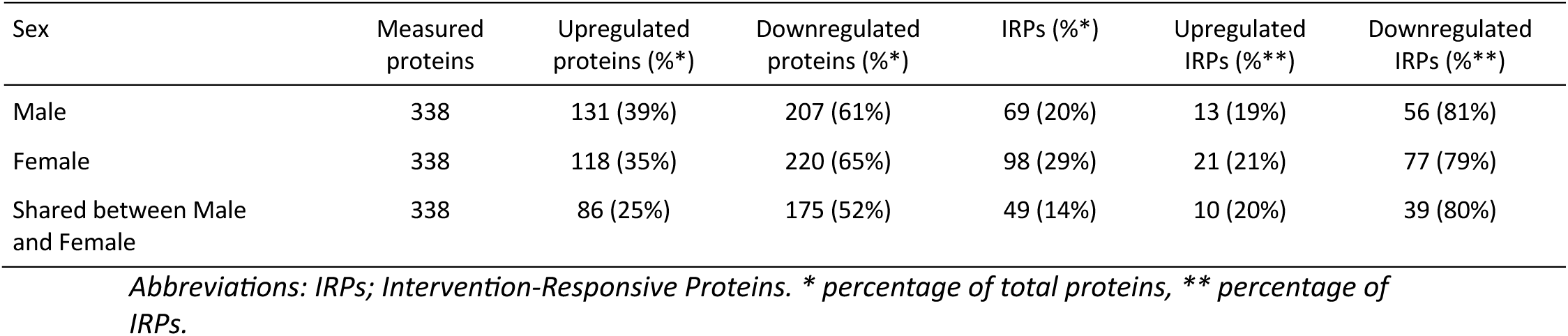
Overview of number of proteins that change as a result of the intervention in male and female participants.

In males, 69 proteins associated significantly with the intervention (56 downregulated, 13 upregulated). A large part of the downregulated proteins is involved in the complement activation (C1R, C2, C3 C4A, C4B, C4BPA, C5, C8B, C8G) and in the blood coagulation cascade (F5, F10, FN1, VTN) (Figure 2, Supplementary Table 4, 5 & 6), whereas the upregulated proteins were largely involved in the adaptive immune response (IGHG2, IGHV1-45, IGHV1-69-2, IGHV3-15, IGHV3-30-3, IGHV3-72, IGKV1-39, TF) and response to stimuli (ALB, CHMP4C, TF) (Supplementary Figure 1).

**Figure 2:**
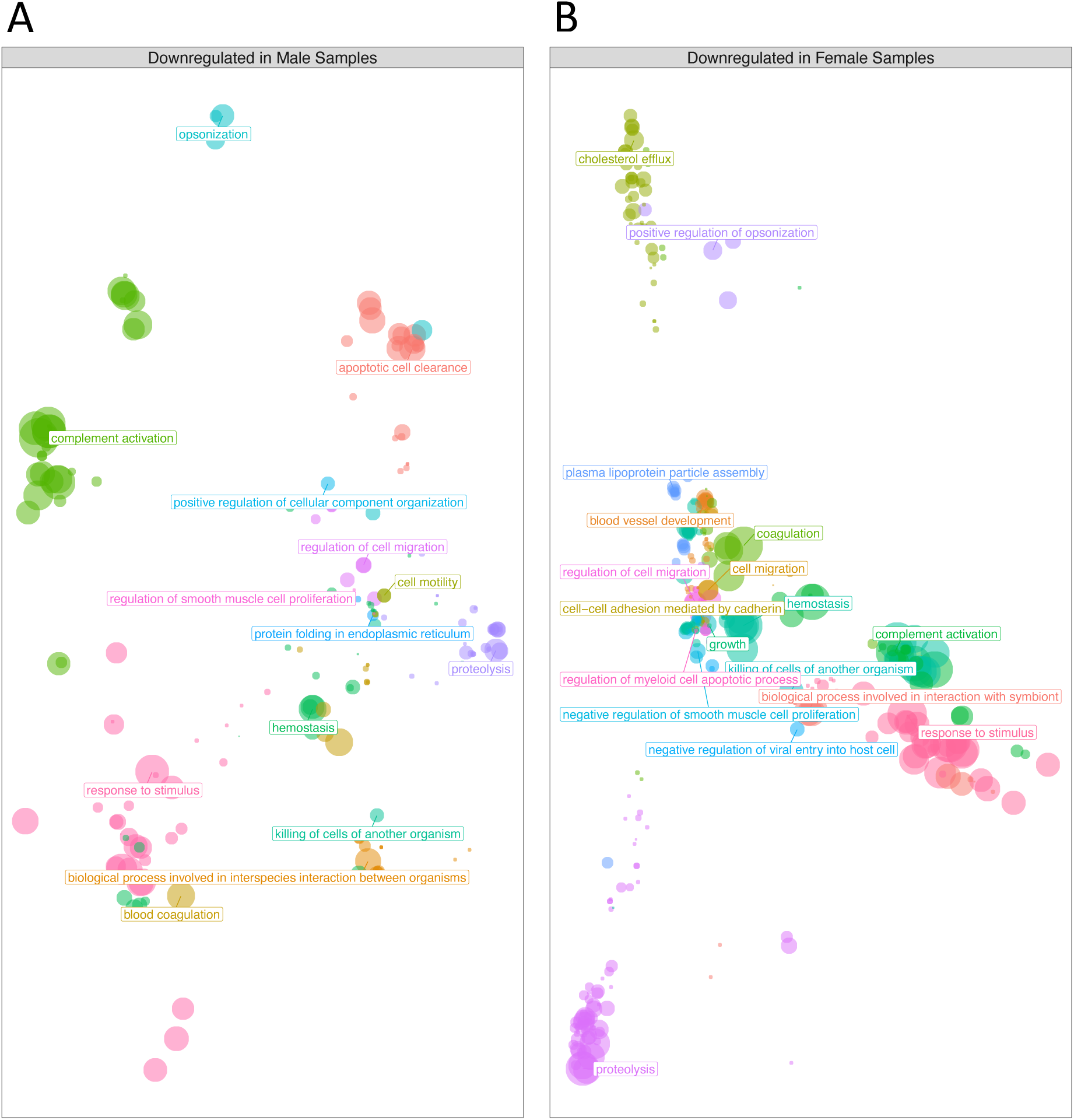
The intervention reduced protein levels involved in the innate immune system. Functional enrichment of significantly downregulated proteins in males (A) and females (B). Enrichment was calculated using a one-sided gene list functional enrichment test. Each dot represents an overrepresented pathway. Dots with the same color represent pathways related to each other. Distance between pathways represent the rela3on between pathways, with closely related pathways (i.e., overlapping genes) plotted near each other. Similar pathways are labeled with their shared parent term. Overrepresented pathways were FDR-adjusted for multiple testing, an adjusted p-value < 0.05 was considered significant.

In females, 98 proteins associated significantly with the intervention (77 downregulated, 21 upregulated) (Figure 1, Table 3). Among the implicated pathways, the blood coagulation cascade (F2, F5, F9, F10, PROS1, SERPIND1, PROZ), plasma lipoprotein particle assembly (APOB, APOC3, APOE) and cholesterol efflux (ADIPOQ, APOB, ACOC, LYVE1, PRG4) showed the strongest overrepresentation of downregulated proteins (Figure 2). Interestingly, the significant downregulation of pathways involved in cholesterol metabolism was specific only to the female participants. Like in the male participants, the upregulated proteins were also involved in the adaptive immune system (Supplementary Figure 1) (IGKV3-20, IGKV3D-15, IGHV1-69-2, IGHV3-30-3). See Supplementary Tables 5 and 6 for a complete list of identified pathways before and after simplification of Gene Ontology terms.

### Downregulated intervention-responsive proteins associate with immune-metabolic health benefits of the intervention

To inquire how the proteomic changes reflected the immune-metabolic health changes, we investigated whether the IRPs associate with the health markers across both time points of the intervention. In males, the intervention responses of all health markers were significantly associated with one or more of 44 (69%) out of the 69 IRPs (subset is shown in Figure 3, all associations are shown in Supplementary Figure 2, Supplementary Table 6). The effects were particularly strong for body composition, serum markers of fat metabolism and inflammation. Even though most of the 44 IRPs were significantly associated with multiple health markers, six of them stand out: PRG4, APOL1, VTN, C3, FN1 and BCHE, as they were significantly associated with at least 7 out of 14 health markers, with PRG4 significantly associated with 10 of the health markers. The direction of all associations was positive, meaning that higher levels of these proteins were associated with higher levels of all health markers, except HDL cholesterol level and size for which the opposite was the case. Notably, all IRPs that associated most prominently to the health marker changes were downregulated by the intervention (Figure 1, Figure 3, Supplementary Figure 2). Interestingly, many of the IRPs were significantly associated with GlycA, but not to fasting CRP.

**Figure 3:**
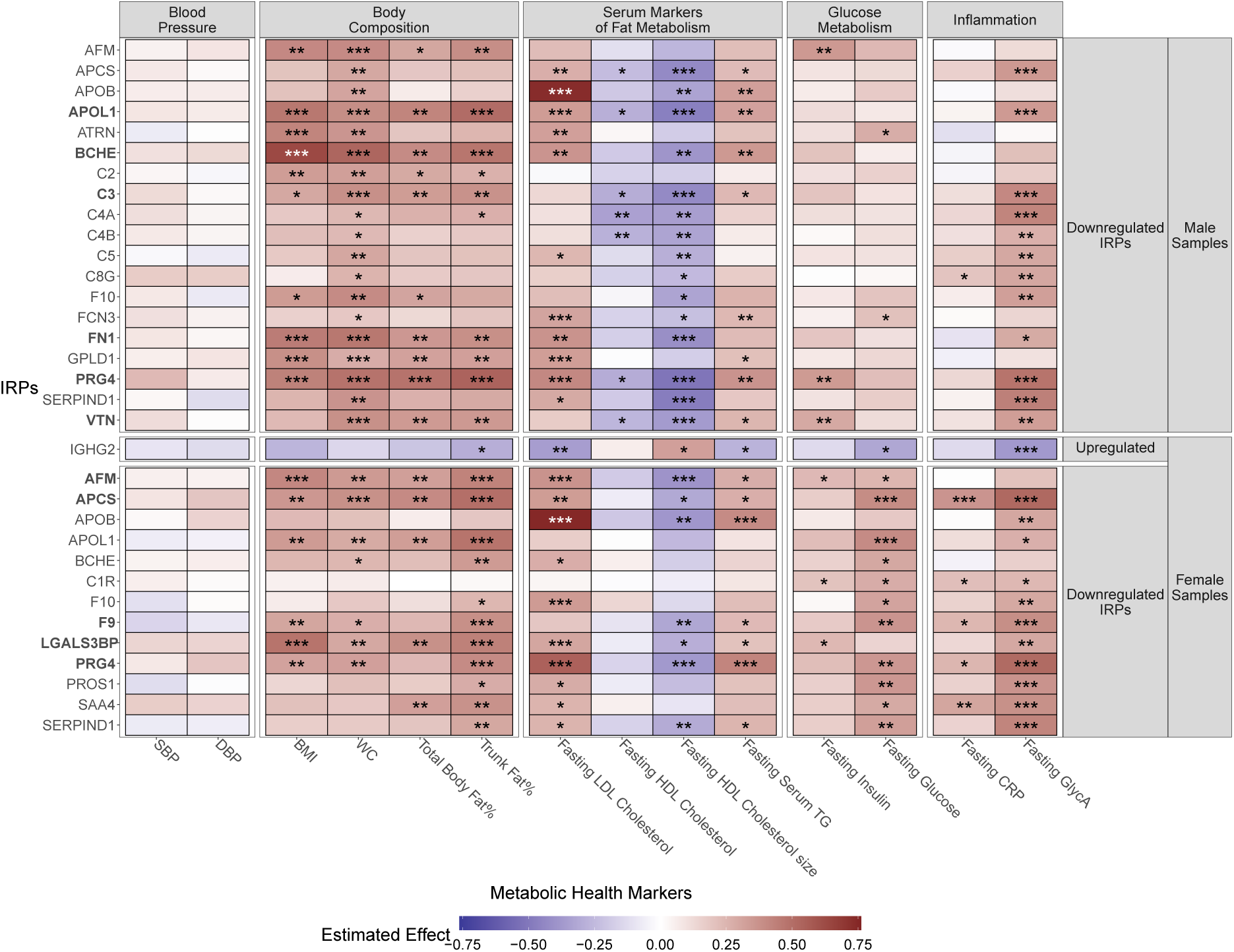
Downregulated intervention response proteins associate with the intervention effects of immune-metabolic health markers. Associations between IRP (intervention response proteins), blood pressure, body composition, serum markers of fat metabolism, glucose metabolism and inflammation. Associations were performed using a two-sided linear mixed model. IRPs are plotted as rows, and health markers in columns (only IPRs that are significantly associated with at least four health markers are shown). Bold gene symbols represent IRPs that are significantly associated to more than half of the measured health markers. Colors indicate the estimated effect between the protein levels and the metabolic health marker levels; blue, a negative effect, white, no effect, red, a positive effect. P-values were adjusted for multiple testing using the false discovery rate. The significance level was indicated by the asterisks: * FDR < 0.05, ** FDR < 0.01, *** FDR < 0.001. Abbreviations: SBP; systolic blood pressure, DBP; diastolic blood pressure, BMI; body mass index, WC; waist circumference, LDL; low-density lipoprotein, HLD; high-density lipoprotein, TG; triglycideres, CRP; C-reactive protein, GlycA; alpha-1-acid glycoprotein.

In females, of the 98 IRPs, 59 were significantly associated with at least one health marker (Figure 3, Supplementary Figure 2, Supplementary Table 7). The effect sizes of these associations were especially strong for body composition, fat metabolism, glucose metabolism and inflammation related health markers. Similar to the results in males, the effect sizes between the downregulated IRPs and the health markers were considerably larger than those of the upregulated IRPs. Additionally, in females, five of the downregulated IRPs captured the improved health effect by at least half of the health markers: especially the association of APCS, PRG4, LGALS3BP, AFM and F9 to body composition, fasting LDL cholesterol, HDL cholesterol size, glucose and GlycA showed a large effect size. Notably, APCS was strongly positively associated with CRP as well.

In general, the pattern of significant associations was similar in male and female participants, with shared significant associations across multiple health marker categories. Most interestingly, PRG4 was captured in both female and male participants. Conversely, there were several differences between male and female associations particular with fasting insulin and glucose.

To better understand the differences in the association of IRPs and health markers, we performed an enrichment analysis of the significantly health marker-associated IRPs to each of the different health marker groups (body composition, serum markers of fat metabolism, glucose metabolism and inflammation). Even though many of the overrepresented pathways overlapped, this still revealed different functional processes that associate with the intervention responses of the health markers. For example, in males, only the proteins associated with serum markers of fat metabolism were significantly overrepresented for triglyceride metabolic process and very-low-density lipoprotein particle assembly, whereas solely the proteins associated with inflammation were overrepresented for liver regeneration & positive regulation of vascular endothelial growth factor production (Supplementary Figure 3). Similarly, in females, the IRPs associated with serum markers of fat metabolism were overrepresented for endocytosis, while the IRPs significantly associated with serum markers of glucose metabolism were overrepresented for cell motility (Supplementary Figure 4). Overall, both the up-and downregulation of blood-based health marker associated IRPs corresponded with favourable changes in health markers, i.e., matching health benefits due to the intervention.

### The intervention responsive proteins associate with the immune-metabolic health status at baseline

Since we observed that the intervention-induced changes in most of the IRPs captured the changes in different aspects of health, we wondered whether the baseline levels of part of the IRPs can also be indicative of the baseline health status as measured by the health markers. We investigated this by associating baseline health marker to baseline IRP levels. Even though the number of significant associations between the baseline protein levels and baseline health marker levels were considerably lower at baseline than when considering the intervention effect, there were some proteins that showed overlapping associations (Supplementary Figure 5, Supplementary Table 8). In males, the baseline levels of 16 IRPs were mainly significantly associated with baseline levels of GlycA and LDL cholesterol. In females, the baseline levels of 33 IRPs were mainly significantly associated with baseline levels of GlycA, LDL cholesterol and triglycerides.

Interestingly, some of the IRPs that best tracked the changes in the health markers, also best captured the baseline health status. This effect was especially strong in the female participants where PRG4 and APCS were significantly associated with 8 and 6 health markers at baseline, respectively. Especially PRG4 showed large effect sizes with trunk fat %, fasting LDL cholesterol, HDL cholesterol size, serum TG, CRP and GlycA at baseline. These results indicate that a subselection of the proteins responding to the intervention not only track the health changes across the intervention but are also reflective of the baseline health status, particularly in female participants.

### Part of the Intervention responsive proteome associates with the blood, subcutaneous adipose tissue and muscle transcriptomic responses

To better understand how the physiological state of metabolic tissues may influence the blood proteome, we investigated to what extend the intervention responsive proteins in the circulation correspond to the responses of the blood, muscle and subcutaneous adipose tissue (SAT) transcriptome in addition to that in blood itself.

In males, five (7%) of the 69 IRPs were significantly associated with the expression level changes of their corresponding genes in the blood: PROSA, TNXB, HSP90B1, ITIH4 & ERN1, the latter three (4%) with a positive direction (Figure 4A & C). Interestingly, twelve of the IRPs (17%) were significantly associated with the matching SAT gene expression level responses. Nine (13%) of these were significantly positively associated with the SAT expression level (C2, C4B, FN1, IGFALS, IGHG2, LYVE1, PRG4, SERPINF1, TNXB) and three (4%) negatively (C5, HSPA5, IGKV3-20). Additionally, eleven IRPs (16%) were significantly associated with the matched gene expression level in muscle; three (4%) positively (C3, IGKV3-20, SERPINF1) and eight (12%) negatively (ANG, C2, C5, C8G, GPLD1, GSN, HSPA5, IGFALS).

**Figure 4:**
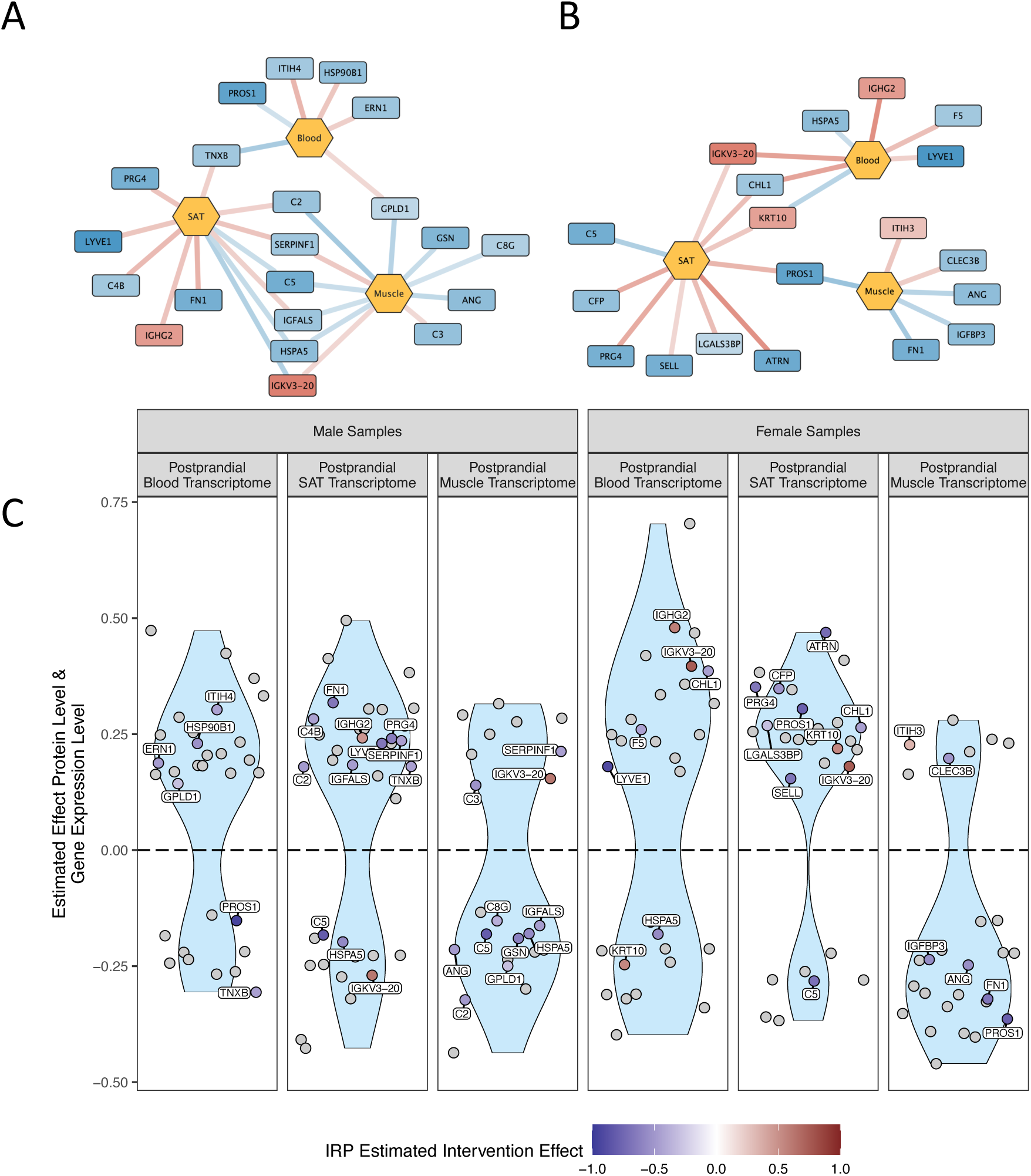
Blood intervention response protein (IRP) levels are significantly associated to gene expression levels in blood, subcutaneous adipose tissue and muscle tissue. (A, B) Network of male (A) and female (B) IRPs protein level and gene expression level. Associations were calculated using a linear mixed model. Red nodes represent upregulated IRPs, blue nodes represent downregulated IRPs. Blue edges represent a negative association between the protein level and the RNA level. Red edges represent a positive association between the protein level and the RNA level. (C) Violin plot of the significant associations between protein levels and matching gene expression levels. Association estimates between the protein level and gene expression level are plotted on the y-axis. Blue points represent downregulated IRPs, red points represent upregulated IRPs. Grey points represent proteins that are significantly associated to the gene expression level but are not significantly changed by the intervention. Abbreviations: IRP: intervention responding protein, SAT: subcutaneous adipose tissue.

In females, seven (7%) of the 98 IRPs were significantly associated with the corresponding gene expression in blood, five (5%) positively (CHL1, F5, IGHG2, IGKV3-20, LYVE1) and two (2%) negatively (HSPA5, KRT10) (Figure 4B & C). Similar to the results in males, the IRP protein levels were strongest associated with the SAT expression levels, with nine (9%) significant positive associations (ATRN, CFP, CHL1, IGKV3-20, KRT10, LGALS3BP, PRG4, PROS1, SELL), and one (1%) significant negative association (C5). In relation to the muscle expression levels, two IRPs (2%) were significantly positively associated (CLEC3B, ITIH3) and four (4%) negatively associated (ANG, FN1, IGFBP3, PROS1).

Out of the three tissues, the IRPs showed the largest number of significant associations with the corresponding gene expression levels in SAT (Figure 4C). Moreover, most of these significant associations were positive showing that the intervention had the same direction of effect on the tissue transcriptome as the circulating proteome, which was not the case in muscle. Lastly, this pattern was also true for the proteins that did not significantly respond to the intervention but were significantly associated with the gene expression level in one or more of the three tissues (Figure 4C, grey dots). These patterns were similar in both sexes. These results indicate that the IRPs with a positive association to expression of the corresponding genes in either the adipose and/or muscle tissue could have been produced in one or both tissues and secreted into the blood stream. This is even more probable if there is no association or a negative association between the IRP protein level and the expression level of the matched gene in blood, such as PRG4 and TNXB.

### Part of the IRPs are mapped to adipose and muscle tissue in the Human Protein Atlas

To investigate whether the IRPs with a significantly positive association between the protein level in blood and the gene expression level in SAT and/or muscle tissue (denoted here as ‘gene IRPs’ (gIRPs)) were also known to be expressed in these tissues, we mapped the gIRPs to the Human Protein Atlas ^27^. In males, out of the nine SAT associated gIRPs, seven were mapped to subcutaneous and/or visceral adipose tissue (C2, FN1, IGHG2, LYVE1, PRG4, SERPINF1, TNXB) (Supplementary Table 9). Additionally, two of the three gIRPs associated with muscle, were mapped to muscle tissue in the Human protein atlas (IGKV3-20, SERPINF1) (Supplementary Table 9). In females, the results were similar. Out of the nine SAT associated gIRPs, four were mapped to adipose tissue in the Human Protein Atlas (CFP, IGKV3-20, PRG4, SELL). Of the two gIRPs associated with muscle, ITIH3 was mapped to muscle tissue. Out of the SAT and muscle gIRPs, C2, FN1, SERPINF1 and ITIH3 were known to be secreted to blood (Supplementary Table 9 & 10). These results further strengthen the hypothesis that some of the IRPs were not expressed in the blood but instead originated from a different tissue and were either actively or inactively secreted into the blood.

### IRPs mapped to adipose and muscle tissue can be used as baseline selection tools for future lifestyle interventions

Since part of the 11 gIRPs described above (C2, CFP, FN1, IGHG2, IGKV3-20, ITIH3, LYVE1, PRG4, SELL, SERPINF1, TNXB) were associated to the baseline health status (Supplementary Figure 5) and the health change during the intervention (Figure 3), we explored whether the baseline protein level was linked to a stronger response in the different health markers. We calculated the baseline tertiles and looked at the mean effects of the studied diagnostic health markers per tertile. We found that especially the baseline levels of IGHG2, IGKV3-20, PRG4 and SELL strongly associated to a stronger health benefit during the intervention, reflected by markers of fat metabolism, glucose metabolism and inflammation (Supplementary Figure 6 – 9). For IGHG2 and IGKV3-20, proteins that increased during the intervention, the health benefits were strongest for participants with a low baseline value (tertile 1). Interestingly participants with a high baseline value for PRG4 and SELL, two proteins that decrease during the intervention, (tertile 3) also benefited from the intervention. Overall, the trends were similar between male and female participants, with slightly stronger effects found in females. Stratifying by tertiles had the strongest effects on the benefits of fasting levels of LDL cholesterol, HDL cholesterol size, serum TG, insulin, glucose, GlycA, and to a lesser extent, CRP (Supplementary Figure 6 – 16). These results indicate that specific IRPs could also be used as baseline tools to select participants who would benefit strongly from a lifestyle intervention.

## Discussion

By investigating the effects of a moderate 13-week combined lifestyle intervention on the fasting blood proteome in older adults (Growing Old TOgether (GOTO) trial, registration number GOTNL3301 (https://onderzoekmetmensen.nl/nl/trial/27183)), we have shown significant responses of the blood proteome to a moderate change in lifestyle. Our results emphasize the applicability of circulating proteins as accurate blood-based biomarkers for health improvements in age-related deficits affecting body composition, fat and glucose metabolism and low-grade inflammation. Our data reveal that a considerable part of the significantly responding proteins (IRPs) is unlikely to originate from the blood, but might be produced in intervention-affected tissues, such as muscle (2-4%) and (subcutaneous) adipose tissue (9-13%), and are (inactively) secreted into the blood. Our findings suggest that the blood proteome is a more informative omics-layer than the blood transcriptome to track health interventions.

The 13-week GOTO intervention had a significant effect on the fasting blood proteome in both sexes, with more pronounced effects in female participants. Notably, roughly 80% of the significant IRPs were downregulated. These downregulated IRPs were overrepresented for biological processes involved in cholesterol efflux, proteolysis, smooth muscle cell differentiation, and different aspects of the immune system, especially inflammatory processes. Moreover, many of the (downregulated) IRPs, were significantly associated with the changes in immune-metabolic health markers. Of particular interest is the strong association with fasting alpha-1-acid glycoprotein acetyls (GlycA), a pro-inflammatory biomarker of which elevated levels have been linked to reduced life expectancy ^28^, and in hospitals to the mortality of older adults ^29^, cardiovascular complications in rheumatoid arthritis ^30^, all-cause population mortality ^31,32^ and a wide scale of incident diseases ^33^. However, since the GlycA measurement used in the GOTO trial is a composite marker ^34^, it is difficult pinpoint the mechanistic process underlying a change in GlycA.

In the GOTO trial, a remarkable part of the significantly downregulated IRPs was involved in inflammatory pathways, particularly in the complement system (C1R, C2, C3, C4A, C4B, C4BPA, C5, C8B, C8G) and coagulation (F2, F5, F9, F10, F12, F13B) (Figure 1, Supplementary Table 4, 5 & 6). A high blood level of these proteins can be an indicator of chronic low-grade inflammation. Since these proteins go down in level across the intervention, this could indicate that the GOTO participants reduced their inflammatory levels, including low-grade inflammation. A considerable part of the IRPs (21 in males, 30 in females) were significantly associated with GlycA, including complement system proteins indicative of low-grade inflammation (PRG4, APCS, SERPIND1) (Figure 3, Supplementary Table 7). The proteins that associate with the change in GlycA, could provide help in understanding the underlying process of changes in GlycA. Interestingly, fasting CRP, a well-studied inflammatory marker was weakly associated with the proteome compared to GlycA (Figure 3, Supplementary Figure 2), and was hardly affected by the GOTO intervention (Table 2). These results indicate that GlycA may be a more sensitive marker or early indicator of inflammation as compared to CRP and reveal clearer differences in inflammageing ^33^, something which currently has been largely overlooked ^35^. Since GlycA and several inflammatory-related proteins were downregulated by the GOTO intervention, it can be postulated that the GOTO intervention had a beneficial effect on the participants’ inflammatory levels ^4^.

The effects of a change in lifestyle on the fasting blood proteome has not been widely studied. The NUTRIAGINGPROT study investigated the effects of a 6-week nutritional intervention (high protein diet) supplemented with 8 weeks of resistance training on plasma protein levels in 134 older adults (62 males 72 females) ^11^. Participants of the high protein diet group within the NUTRIAGINGPROT study significantly increased their muscle mass, while reducing their body fat %. Within the GOTO trial we also observed a reduction of body fat % as a result of the intervention as well as a reduction of absolute muscle mass, but an increase of muscle mass relative to total mass, suggesting that the GOTO intervention successfully improved the individual’s health state ^20^. In addition, similarly to the GOTO trial, the NUTRIAGINGPROT study identified proteins (14 in total) involved in coagulation, innate immune system and lipid transport. Five of these proteins were significantly downregulated in both studies (F5, F12, GSN, IGFBP3, VTN, Figure 1, Supplementary Table 4) ^11^. Conversely, one protein was downregulated in GOTO and upregulated in the NUTRIAGINGPROT study (CNDP1). The different effect on CNDP1 might be attributed to the change in diet, since CNDP1 only significantly changed in the high protein group, while the GOTO participants reduced their caloric intake. Overall, these results highlight the impact of interventions within humans and indicate the possibility of a largely shared response mechanism that leads to increased health benefits of these type of interventions.

One of the tissues that plays a large role in age-related metabolic dysfunction is adipose tissue ^36^. Previously, we reported that the postprandial blood transcriptome hardly responded to the GOTO intervention, but the transcriptomes of postprandial subcutaneous adipose tissue (SAT) and muscle tissue responded significantly ^16^. Additionally, the expression levels of SAT best captured the immune-metabolic health changes ^16^. This is in line with the intervention itself as it increased physical activity by 12.5% as well as reduced caloric intake by 12.5% in the participants. Since the fasting blood proteome did respond significantly to the GOTO intervention and was strongly associated with immune-metabolic health markers, it is likely that the majority of the IRPs that we have measured in the blood do not originate from transcriptionally and translationally active cells in the blood itself but are rather secreted from other tissue. We showed that ten of the gIRPs associated with adipose tissue, were mapped to adipose tissue in the Human Protein Atlas ^27^ (C2, CFP, FN1, IGHG2, IGKV3-20, LYVE1, PRG4, SELL, SERPINF1, TNXB) and part of them were known to be secreted into the blood (C2, FN1, SERPINF1) (Supplementary Table 9 & 10). A large part of these proteins are involved in inflammatory or ageing related processes. C2 and CFP are both part of the complement system, which plays a role in multiple processes, including inflammatory processes, and has been linked to ageing and age-related diseases ^37^. Additionally, increased production and activation of complement components in adipose tissue have been linked to increased insulin resistance ^38^. Besides complement components two other proteins are regulators of the complement system: CFP, a precursor of the plasma glycoprotein properdin, is involved in complement activation ^39^, and IGHG2 is predicted to be involved in complement activation. LYVE1, a glycoprotein, is also involved in inflammatory pathways, and has been mostly described as a receptor for the extracellular matrix glycosaminoglycan hyaluronan, an abundant component of soft tissue that facilitates cell migration during wounding and inflammation ^40^. PRG4, one of the proteins associated with the change in GlycA (Figure 3, Supplementary Table 7), PRG4 is an extracellular protein with anti-inflammatory properties and is thought to play a significant role in joint lubrication and inflammatory regulation of synovial macrophages ^41,42^, hence PRG4 could therefore be directly related to individuals’ ability to move. SELL (L-Selectin) is another protein that is involved in the immune system, especially through its roles in lymphoid tissue, and on the surface of monocytes and neutrophils where it allows these immune cells to emigrate from the bloodstream into the inflamed tissue ^43,44^. Even though the Human Protein Atlas has mapped SELL to adipose tissue, it is mostly expressed and found in bone marrow and lymphoid tissues ^27^. Lastly, TNXB (transcribes for the extracellular matrix glycoprotein tenascin-X) has not been linked to age-related processes. TNXB has been found to play a role in the stability and the maintenance of the collagen network ^45^, which might be indicative of muscle remodeling during the GOTO intervention ^16^. Many of the processes described above are age- and/or inflammatory-related. In combination with the fact that most of these proteins were downregulated during the intervention (all except IGHG2), these results could indicate that the participants of the GOTO intervention had a healthier ageing and inflammation profile after the intervention than prior to the intervention.

Two of the proteins that were significantly positively associated with the matched gene expression level in SAT, were also significantly associated with the gene expression level in muscle tissue (IGKV3-20, SERPINF1). IGKV3-20 is part of immunoglobulin light chains that participates in the antigen recognition and has not been specifically linked to ageing or age-related processes ^46^. SERPINF1, has been linked to several rodent models of obesity when expressed in high numbers ^47^. Notably, and similar to the results in GOTO, SERPINF1 levels reduced upon weight loss, as well as upon insulin sensitization ^47^. One of the IRPs was uniquely significantly positively associated with the gene expression level in muscle; ITIH3. ITIH3 is part of the ITIH protein family and has been proposed as a biomarker for multiple diseases, including as an early detection for rheumatoid arthritis ^48^. Additionally, in a study investigating the plasma proteome of healthy participants undergoing a 10-day bed rest, elevated levels of ITIH3 were associated with a larger muscle loss ^49,50^. Even though the participants of the GOTO trial reduced their body fat %, they also had a significant reduction in total lean mass (-0.83 kg and -0.51 kg for male and female participants, respectively) ^4^, which could potentially explain why the ITIH3 levels were significantly upregulated in the blood proteome, albeit only in female participants.

The baseline levels of three gIRPs mapped to the adipose (IGHG2, PRG4, SELL) and IKGV3-20, which were mapped to both adipose and muscle tissue, were indicative of a stronger immune-metabolic health improvement during the intervention (Supplementary Figure 6-9). The health improvements were particularly strong for serum markers of cholesterol and glucose metabolism and inflammation, indicating that these four proteins could function as potential selection criteria for novel lifestyle intervention or as a risk marker for immune-metabolic health.

### Limitations of the GOTO trial

When interpreting the observed results, we must acknowledge that the Growing Old TOgether (GOTO) trial was a personalized lifestyle intervention. This meant that the nutritional guidelines as well as the planned physical activities differed between the participants and may thus have introduced variation in the effect of the intervention. In addition, the protein levels were measured in the fasting state in blood, while the blood, subcutaneous adipose tissue and muscle tissue transcriptomes were sampled 30 minutes after a standardized meal challenge. Hence, the physiological conditions of these tissues are different. Muscle tissue is known to provide glycogen and essential nutrients to the blood stream, to a greater extent than the SAT, and therefore muscle tissue is expected to be more greatly deprived of these nutrients. While the effects of the nutrient challenge and their intervention effect are well characterized in the transcriptome of blood ^51^, these short-term effects in muscle and SAT are unknown. Therefore, while the muscle and SAT are certainly expected to react to the increased nutrient availability following the meal, we do not know whether this is within the 30 minutes time frame of sampling nor how this is precisely reflected in the intervention. Finally, we cannot say whether the blood proteomic changes as results of the GOTO intervention remain for a longer time in the circulation, since we did not collect additional longitudinal biological samples from the GOTO trial participants.

### Concluding remarks

Overall, we investigated the effects of a moderate 13-week combined physical activity and dietary lifestyle intervention on the fasting blood proteome of older adults. We highlighted how especially the reduction of the inflammatory related proteins went hand in hand with immune-metabolic health improvements relevant in counteracting ageing related deficits. Through linking the blood protein levels to the blood, SAT and muscle tissue transcriptome data, we showed that part of the circulating proteome, including FN1, LGALS3BP & PRG4, may serve as a proxy to tissue-specific immune-metabolic health changes, especially to SAT. Lastly, we showed that the baseline levels of four circulating proteins mapped to adipose and/or muscle tissue: IGHG2, PRG4, SELL and IKGV3-20, were indicative of a stronger immune-metabolic health improvement. These results could open up the possibility for large-scale tissue-health oriented studies without the need for a biopsy. The blood proteome therefore seems a valuable and sensitive instrument to monitor response to lifestyle interventions and needs to be further explored in a dynamic context in addition to cohort studies.

## Methods

### GOTO Lifestyle Intervention

The Growing Old TOgether (GOTO) trial, is a 13-week nonrandomized, noncontrolled combined lifestyle intervention with 164 older adult participants, one of whom withdrew due to a knee surgery. A thorough description of the trial design, power calculation, inclusion and exclusion criteria, primary and secondary outcomes, and the handling of compliance to the intervention has been described previously ^4,16^. In short, the GOTO participants underwent a 25% energy reduction comprised through 12.5% decreased caloric intake and 12.5% increased physical activity. Personalized intervention guidelines were prescribed by a dietician and a physiotherapist, in consultation with the participant’s preferences and physical abilities. The dietary guidelines were composed to be as much as possible according to the “Dutch Guidelines for a healthy diet” ^4^. As described in ^16^, 153 out of 164 participants were considered compliant to the intervention, out of these fasting blood proteome measurements were available from 146 participants (70 male, 76 female). Out of these 146 participants, transcriptomic measurements of postprandial blood, subcutaneous adipose tissue (SAT) and muscle tissue were available for a subset of 54 participants (31 male, 23 female).

### Proteomic measurements

Samples were prepared following a modified SP3 protocol ^52^ performed on an Integra Assist plus (Integra), foregoing the peptide cleanup on the second day. Instead, beads were removed after acidification and samples cleaned by using mixed-mode StageTips ^53^. Samples were analyzed on an UltiMate 3000 coupled to an Orbitrap Exploris 480 with FAIMS pro (all Thermo Scientific). Samples were loaded onto a precolumn (PepMap 5 mm cartridge, Thermo Scientific) and reverse flushed onto an in-house packed 30 cm pulled tip column (150 µm inner diameter, filled with 2.7 µm Poroshell EC120 C18, Agilent). Separation took place on a 25 min gradient running 0.1 % formic acid (eluent A) against 80 % acetonitrile, 0.1 % formic acid (eluent B). Gradient started at 6 % B and increased to 35 % over 22 min followed by washing and equilibration to standard conditions, all with a constant flow of 1 µl/min. FAIMS set to - 50 V compensation voltage with inner and outer electrode temperature kept constant at 99.5°C and 85 °C, respectively. The mass spectrometer resolution of both MS1 and MS2 were set to 15k resolution and was running in data independent acquisition mode an 30 % normalized collision energy. The mass range from 380 to 900 m/z was covered in 30 staggered windows of 16 m/z each, resulting in effectively 8 m/z windows after deconvolution using ProteoWizard^54^. For library generation, a pool was generated from all samples and high pH fractionated on an 1 h gradient using an Infinity 1260 LC (Agilent). Resulting 96 fractions were concatenated into a total of 24 samples and analyzed on the identical setup and LC gradient used for sample analysis but with the mass spectrometer operated in Top 18 DDA. MS1 resolution was set to 60k, MS2 resolution to 30k with a dynamic exclusion of 20 s and an isolation window of 1.2 Th. The library was afterwards build using Fragpipe 15.0 and its predefined library building workflow with standard parameters and the Human Uniprot reference proteome including isoforms (downloaded 13.01.2021). The resulting library contained 8781 precursors from 681 proteins. Finally, samples were searched against the library using DIA-NN ^55^ 1.8.1 with reannotation activated and the additional command line “—report-lib-info”.

### Proteomics Preprocessing

Raw MS spectra were processed using DIA-NN (v. 1.8.1) and filtered to ensure that each identified protein had at least 1 unique peptide identifying it. Proteins that had more than 80% missing data were removed. Resulting intensity values were then log_2_ transformed. Missing values were imputed using a random normal distribution with a mean equal to the mean of the 5% proteins with the lowest intensities minus 1.6, and a variance equal to the 80% of the standard deviation of the lowest 5% of intensities. This ensured that all missing values were imputed just under the observed intensity levels. Finally, protein values were Z-score scaled within each batch of samples. All statistical analyses were performed in R (v. 4.2.2).

### Differential protein expression analysis & functional enrichment

To identify significantly changed proteins across the intervention, a linear mixed effect model was applied for each sex separately,

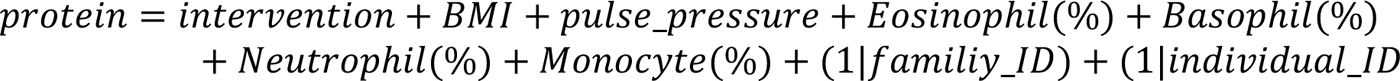

Proteins with an FDR corrected p-value of 0.05 were extracted and were then subjected to functional enrichment using the approach implemented within the gprofiler2 package (v. 0.2.2) ^56,57^. Plots were generated using the ggplot2 (v. 3.4.4) ^58^ and ggrepel packages (v. 0.9.2)^58^.

To ensure we highlight all the functions that may be represented by the identified proteins we opted to not include the measured background within the functional enrichment but used the standard global background. To visualize the results, we used the rrvgo package (v. 1.10.0) ^59^, which groups functional terms based on their semantic similarity index. Similarity threshold of 0.975 was used.

### Diagnostic measurements

All measurements were performed in fasting serum collected through venipuncture. Insulin was measured using a Immulite 2000 xPi (Siemens, Eschborn, Germany). Fasting high-density lipoprotein (HDL) cholesterol and fasting alpha-1-acid glycoprotein (GlycA) levels were measured using the previously described Nightingale platform ^60^. Complete methods of diagnostic measurements are described in van de Rest et al. ^4^.

### DEXA measurements

Method for the Dual-energy X-ray absorptiometry (DEXA) measurements is described in Beekman et al. ^20^. In short: whole-body DEXA (Discovery A, Hologic Inc., Bedford, MA, USA) was used to measure eleven body composition components, including whole-body fat mass and trunk fat mass. Whole-body fat% was calculated by dividing whole-body fat mass by whole-body mass. Trunk fat % was calculated by dividing the trunk fat mass by the total trunk mass.

### Health marker baseline comparisons and intervention effect

The significance level of the difference between the baseline age and baseline health marker levels between male and female participants was calculated using an unpaired t-test. The p-value was subsequently adjusted for multiple testing using the false discovery rate. The intervention effect was calculated using a linear mixed model: health marker = intervention + age at baseline *health maker = intervention + baseline age + (1|familiy_ID) + (1|individual_ID)*. The significance level was subsequently adjusted for multiple testing using the false discovery rate.

### Association analysis of the proteome to health markers

Health markers for each participant were measured both at the start of the intervention (baseline) and after the intervention (post intervention), as described in van de Rest et al. ^4^. Prior to the association analysis, both the protein levels and the health marker levels were Z-scored. The associations between the baseline protein levels and baseline health marker levels were studied using a linear model with age as fixed effect: *protein ∼ health marker + age at baseline* (implemented with the *lm* function of the R package stats (v. 4.2.2)). To explore the relation of the intervention effects on proteome and health markers, associations were investigated across the two time points of the intervention (baseline and post intervention) with a linear mixed model with age at baseline as fixed effect and person ID as a random effect: *protein ∼ health marker + age at baseline + (1|person id)* (implemented with the function *lmer* of the R package lmerTest (version 3.1-3)). P-values were adjusted for multiple testing using the false discovery rate (FDR).

### RNA isolation and sequencing

The full description of the RNA isolation and sequencing has been described in Bogaards et al. ^16^ and Gehrmann et al. ^51^. In short, libraries were prepared using Illumina TruSeq version 2 library preparation kits. Data processing was performed the in-house BIOPET Gentrap pipeline^61^. The following steps were part of the data processing: low-quality trimming using sickle version 12.00. Cutadapt version 1.1 was used to perform the adapter clipping. The reads were aligned to GRCh37, while masking for SNPs common in the Dutch population (GoNL ^62^ MAF > 0.01), using STAR version 2.3.0e. Picard version 2.4.1. was used to perform sam-to-bam conversion and sorting. Read quantification was performed using htseq-count version 0.6.1.p1 using Ensemble gene annotations version 86 for gene definitions. In postprandial blood, the sequencing resulted in an average of 37.2 million reads per sample, 97% ( + − 0.4%) of which were mapped. In postprandial SAT, samples had an average of 11.4 million sequenced reads, 95% ( + − 1.6%) of which were mapped. In postprandial muscle, an average of 36.9 million sequence reads per sample, 98% (+ −0.4%) of which were mapped. Transcriptomics data was available of a subset of the GOTO participants: 88 (45 male, 43 female) participants with postprandial blood measurements, 78 (38 male, 40 female) with postprandial SAT measurements, 82 (48 male, 34 female) with postprandial muscle measurements. A subset of 54 participants (31 male, 23 female) had transcriptomic measurements in all three tissues.

### Associating the blood, subcutaneous adipose tissue and muscle tissue transcriptome response to the observed proteome changes

To investigate the relationship between the protein level and the transcriptomic level of the corresponding gene in blood, SAT and muscle tissue, a linear model was used: *protein level ∼ blood expression level + SAT expression level + muscle expression level* (implemented with the *lm* base function of R). Notably, all three transcriptomic levels were added to the same model. Prior to the estimated effects calculation, the proteome and transcriptome measurements were Z-scored. An FDR-adjusted p-value below 0.1 was considered significant.

## Data Availability Statement

The data that support the findings of this study are accessible upon request to the corresponding author. The data are not publicly available due to privacy or ethical restrictions.

## Author Contributions

P.E. Slagboom and C.P.G.M. de Groot designed the study. M. Beekman collected and curated the health data. N. Lakenberg and E. Suchiman generated the transcriptome data. J.-W. Lackmann and S. Müller generated the proteome data. F.A. Bogaards, P. Antczak, T. Gehrmann, R.-U. Müller, M.J.T. Reinders and P.E. Slagboom designed the data analysis approach. F.A. Bogaards and P. Antczak performed the analysis and F.A. Bogaards, P. Antczak, J. Deelen, R-U. Müller, M.J.T. Reinders and P.E. Slagboom performed the research and interpreted the data. All authors were involved in drafting and revising the manuscript.

## Ethics Statement

The Medical Ethical Committee of the Leiden University Medical Center approved the study (P11.187) and all participants signed a written informed consent. All experiments were performed in accordance with relevant and approved guidelines and regulations. This trial was registered at the Dutch Trial Register (https://onderzoekmetmensen.nl/en/trial/27183) as NL3301 and can also be found at the international clinical trials registry platform as NL-OMON27183.

## Funding and Acknowledgements

This work was funded by the Netherlands Consortium for Healthy Ageing (NWO grant 050-060-810, P.E.S.) and the ZonMw Project VOILA (ZonMW 457001001, P.E.S.). T.G. was funded through the European Research Council (starting grant Lacto-Be 852600). R.U.M. was supported by the Ministry of Science North Rhine-Westphalia (Nachwuchsgruppen.NRW 2015-2021), the German Research Foundation (DFG DI 1501/9, DFG MU 3629/6-1), the Marga and Walter Boll Foundation, and the PKD Foundation. P.A. and R.U.M. received support from the joergbernards-Stiftung as well as Köln Fortune and CECAD. We acknowledge support for the Article Processing Charge from the DFG (German Research Foundation, 491454339). This work was supported via DFG large invest grants (DFG Großgeräteantrag INST 1856/71-1 FUGG & INST 216/1070-1 FUGG). The funding agencies had no role in the design and conduct of the trial; collection, management, analysis, and interpretation of the data; and preparation, review, or approval of the manuscript. The authors would like to express their gratitude to all participants of the GOTO trial who did their very best to adhere to the intervention guidelines and underwent all measurements.

