## Supplementary figures and images for "Blood Proteomic Biomarkers indicate reduced system and tissue level inflammation responses to The GOTO Lifestyle Intervention in Older Adults"

### Supplementary Figure 1

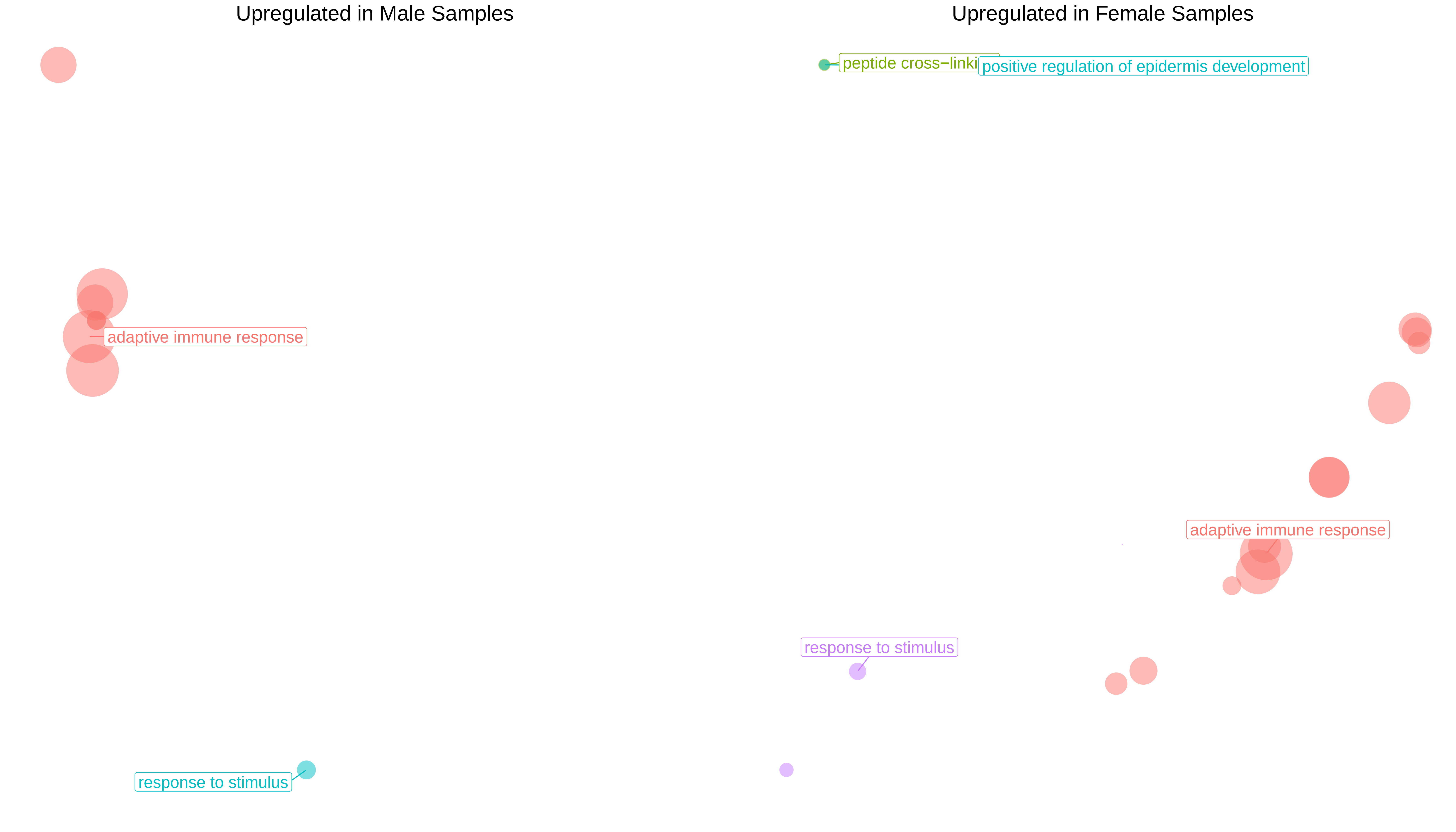

### Supplementary Figure 2

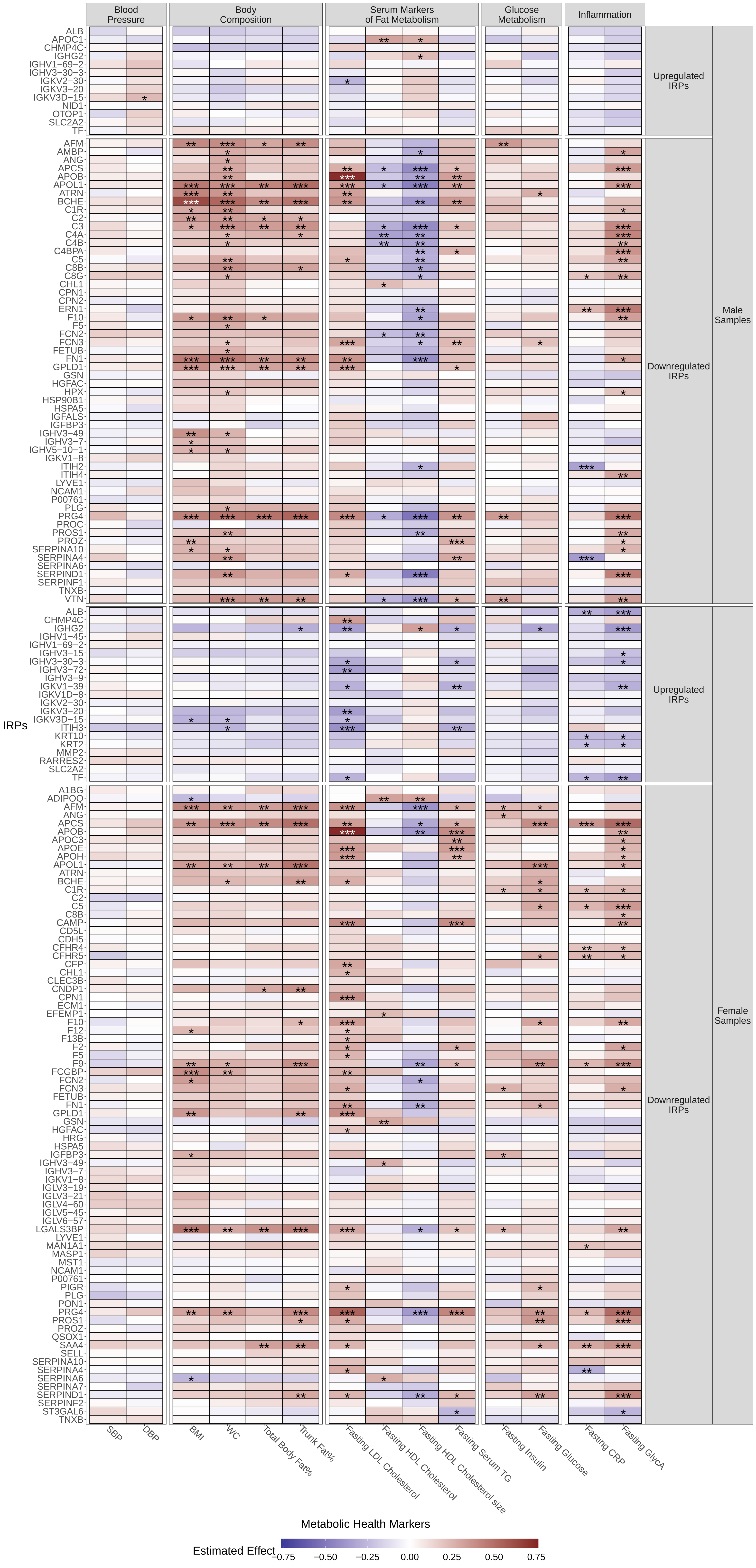

### Supplementary Figure 3

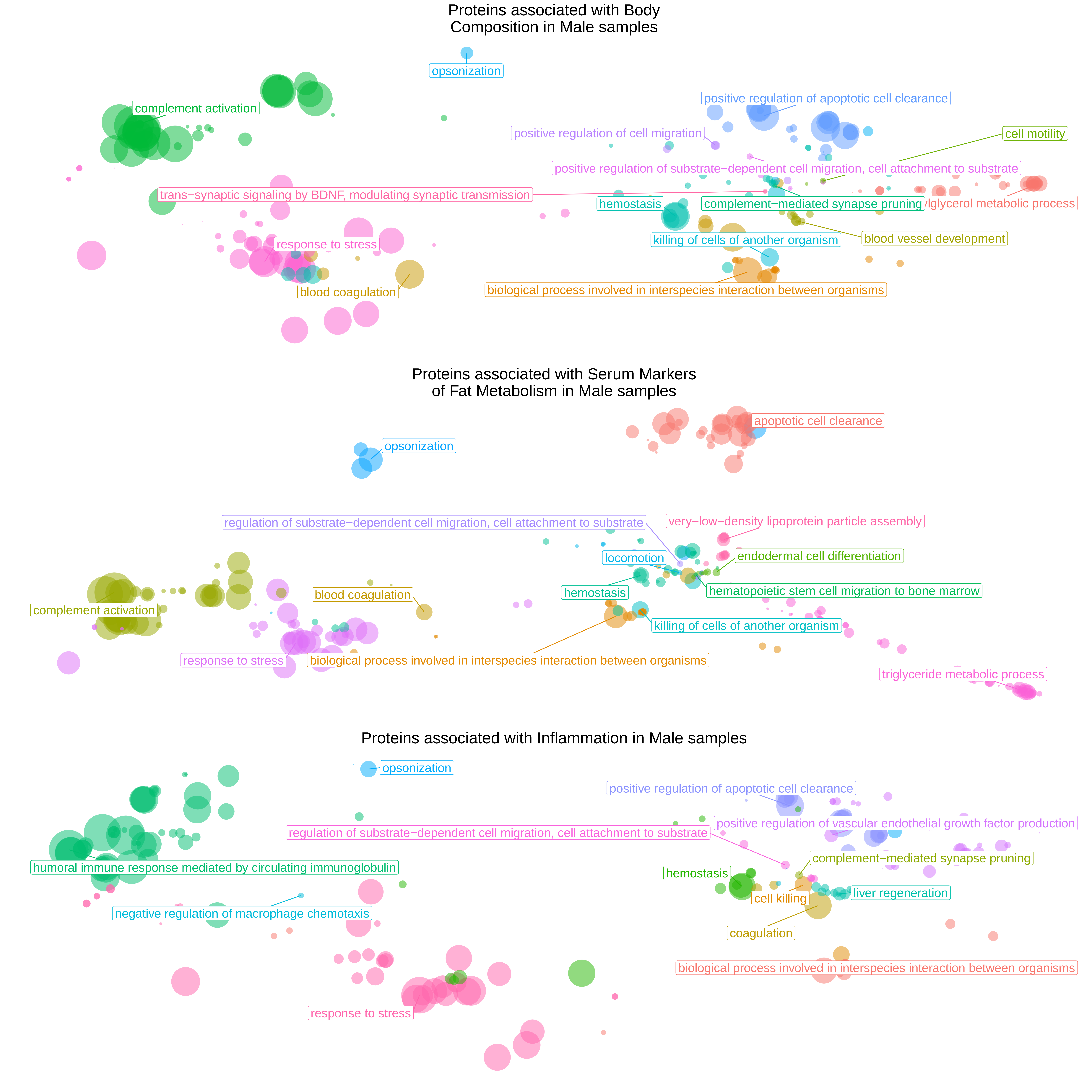

### Supplementary Figure 4

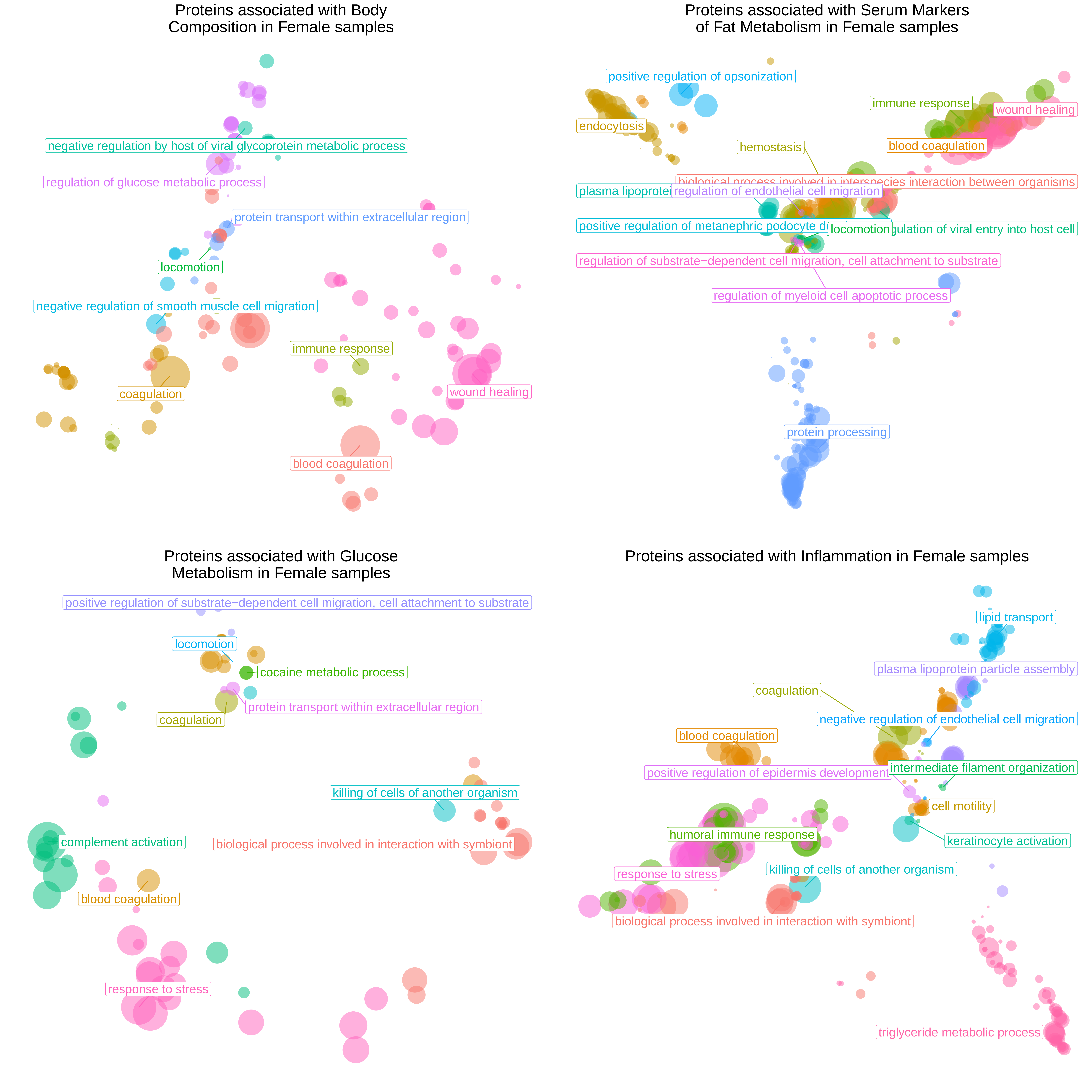

### Supplementary Figure 5

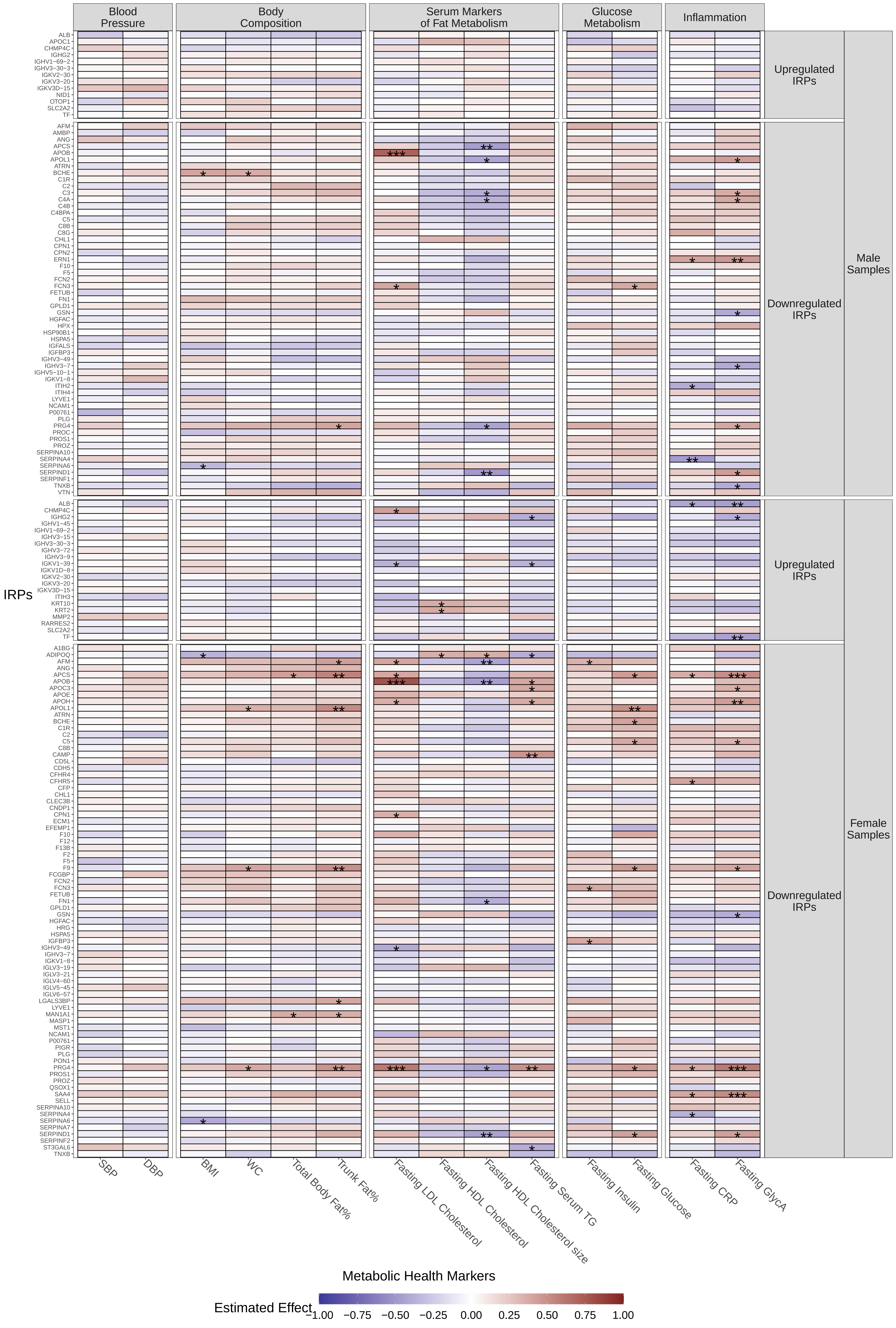

### Supplementary Figure 6

#### Tertiles using IGHG2 baseline values

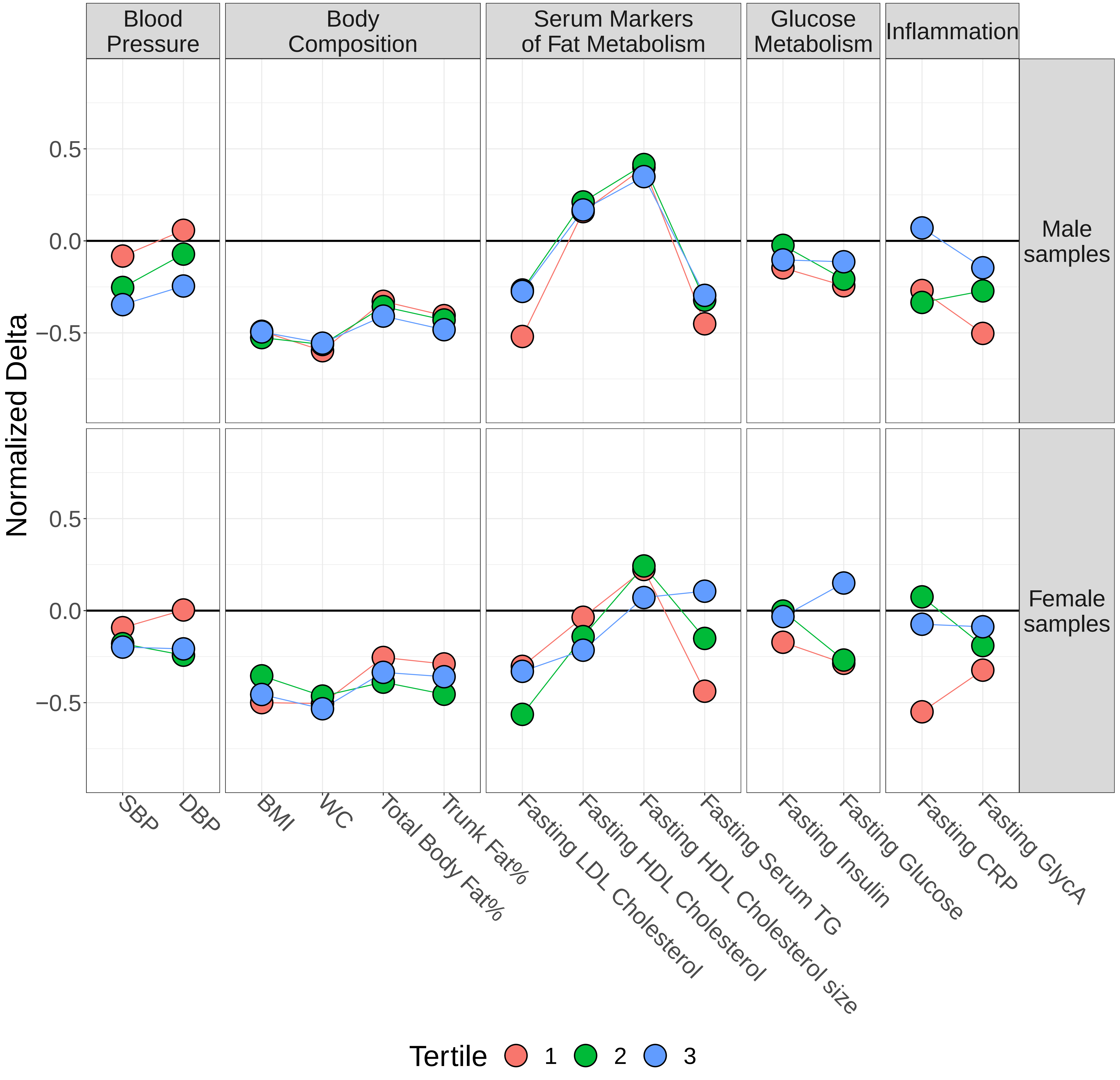

### Supplementary Figure 7

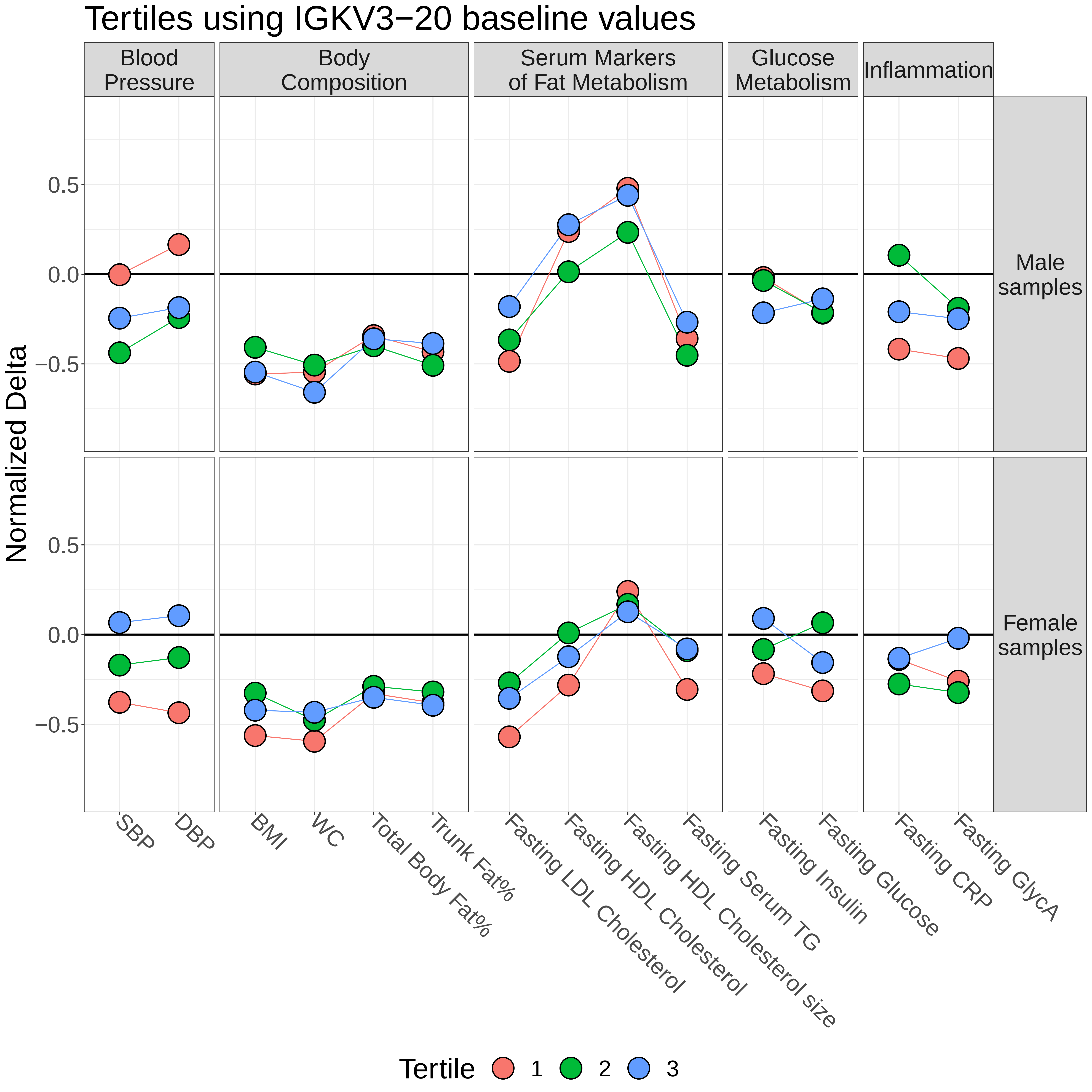

### Supplementary Figure 8

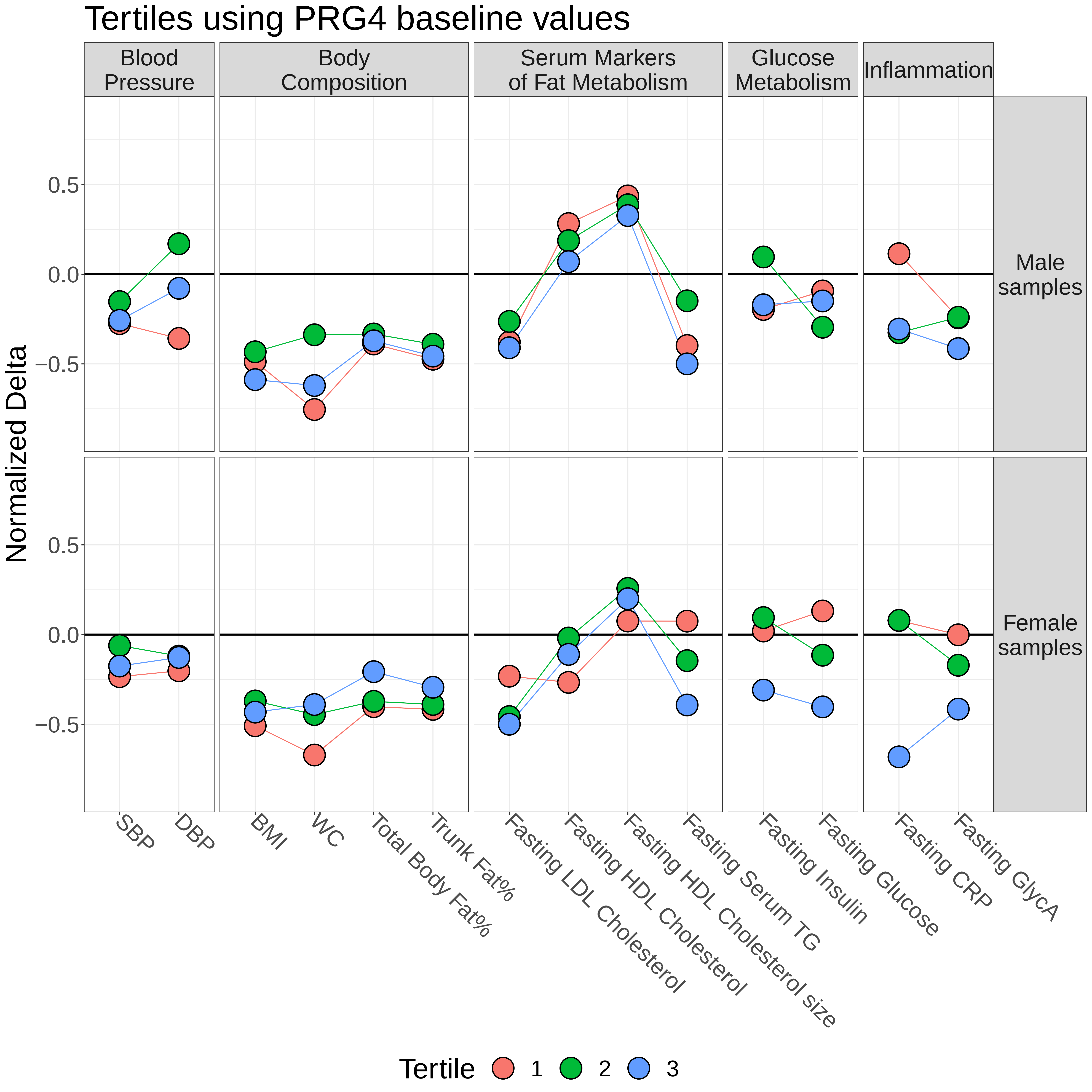

### Supplementary Figure 9

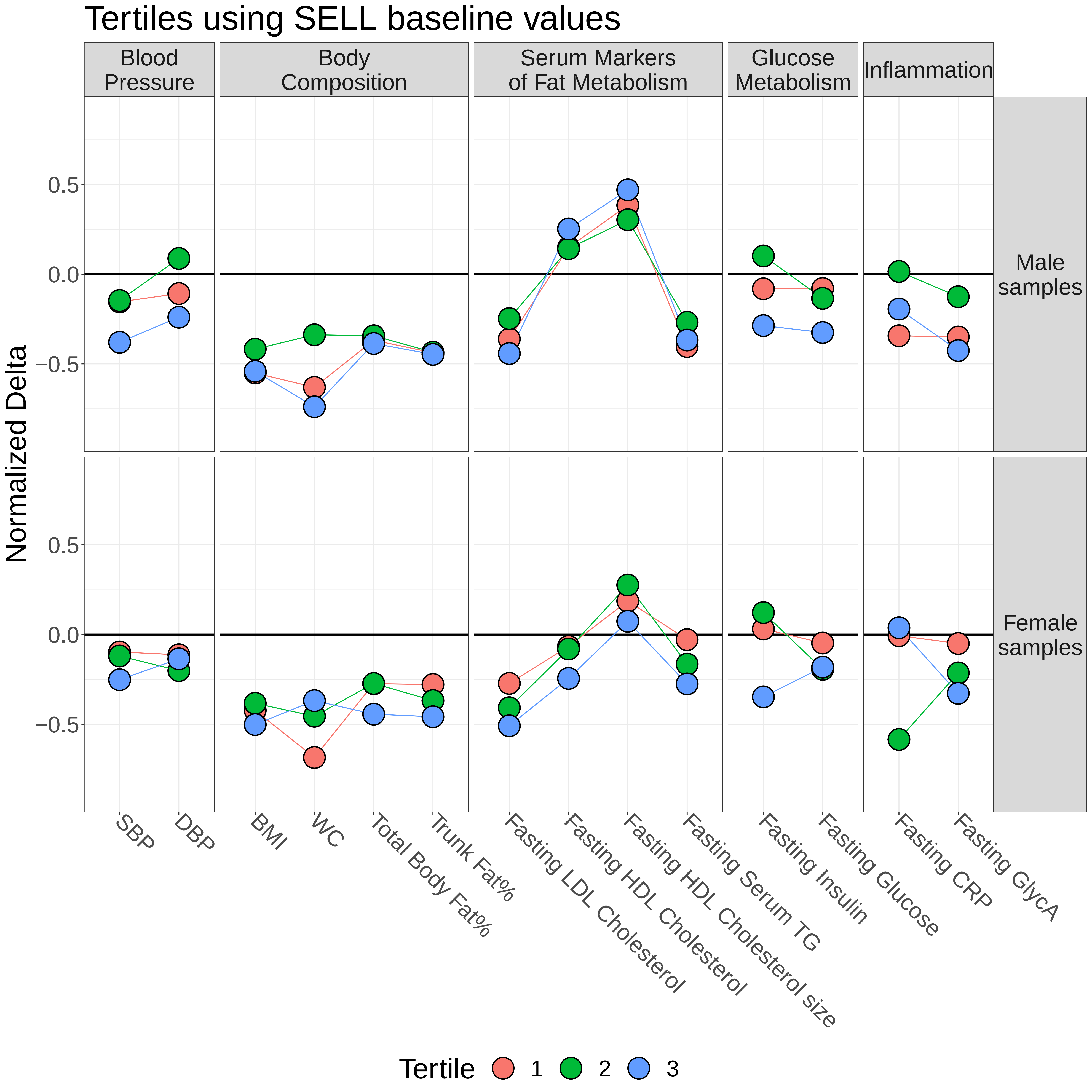

### Supplementary Figure 10

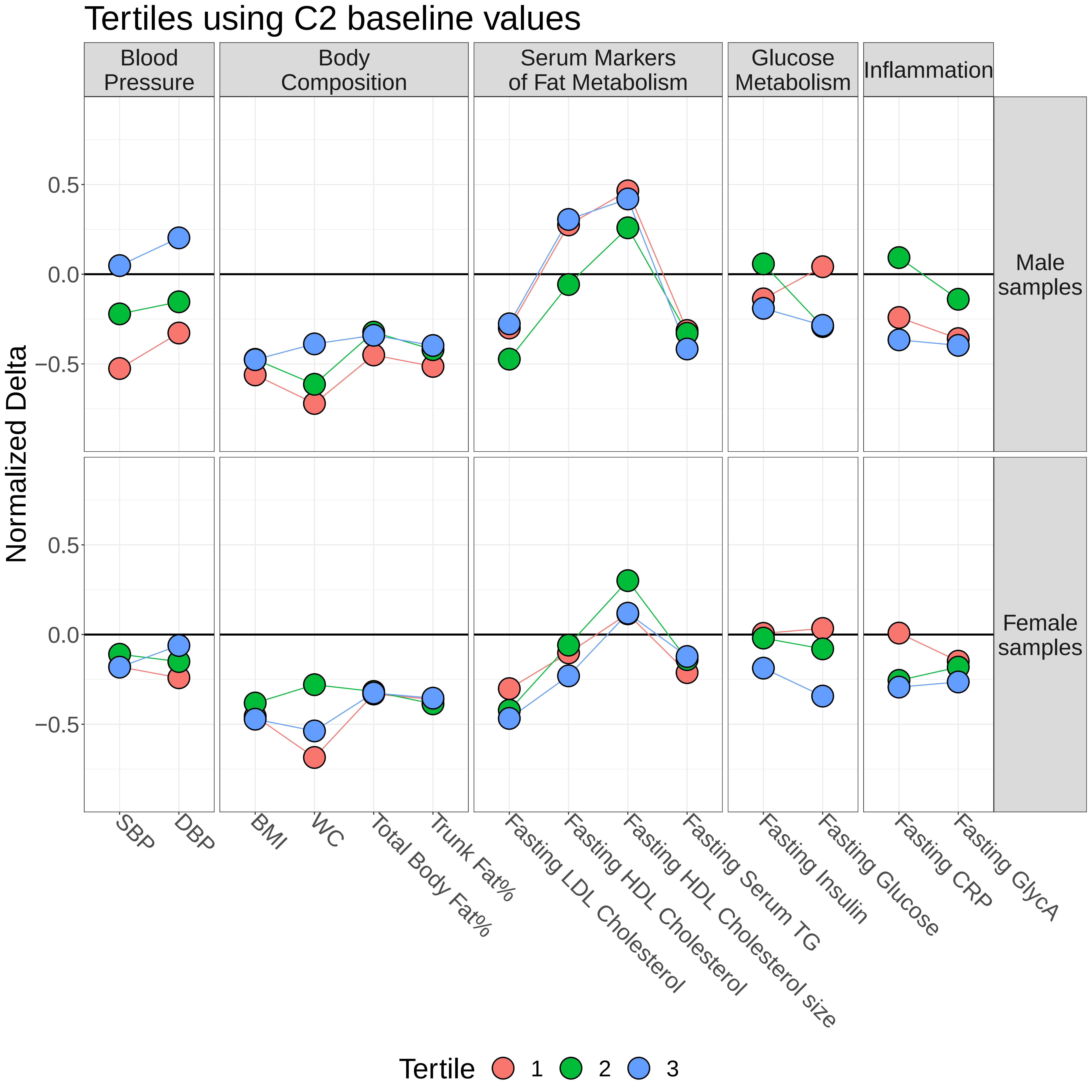

### Supplementary Figure 11

#### Tertiles using CFP baseline values

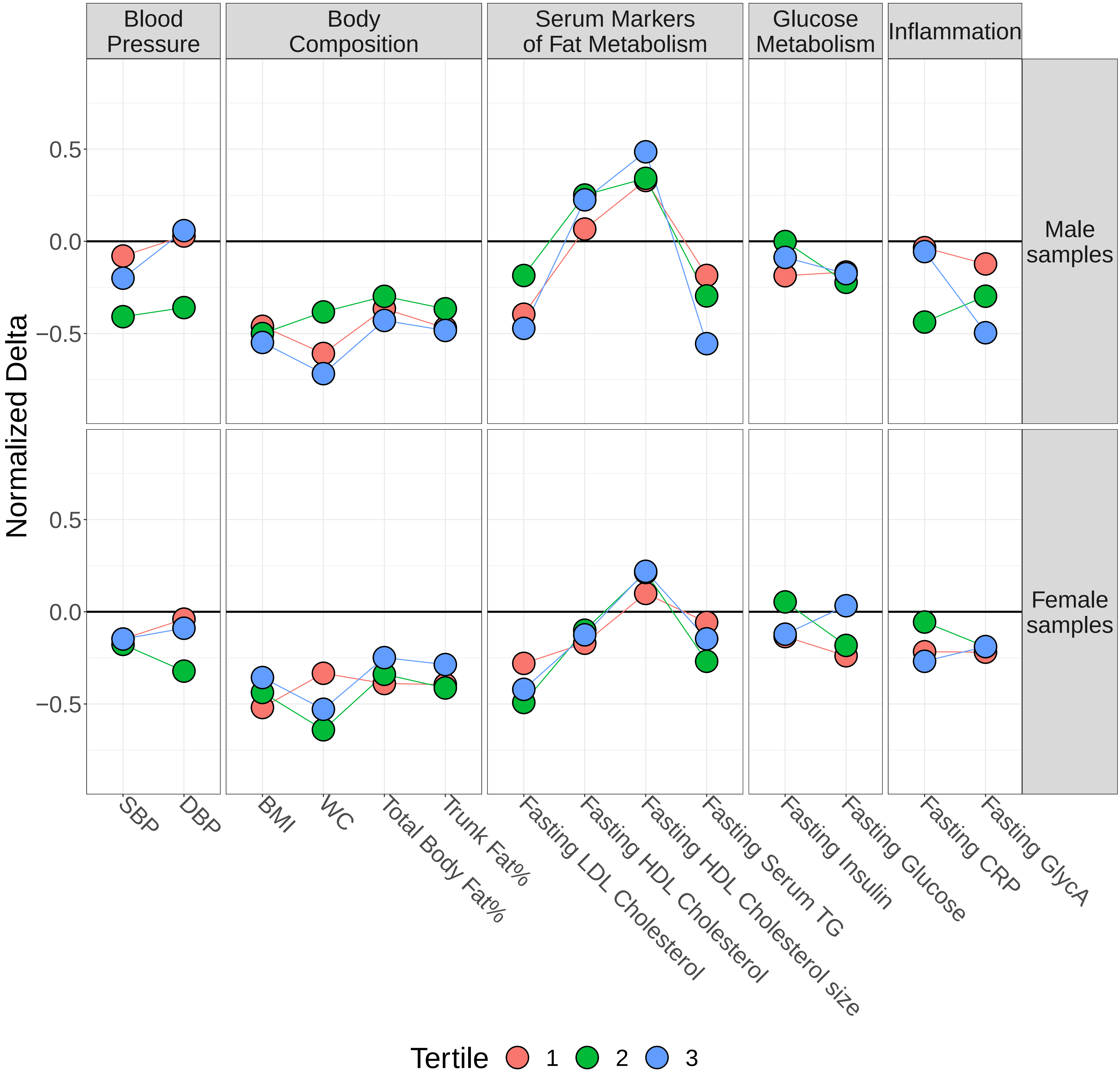

### Supplementary Figure 12

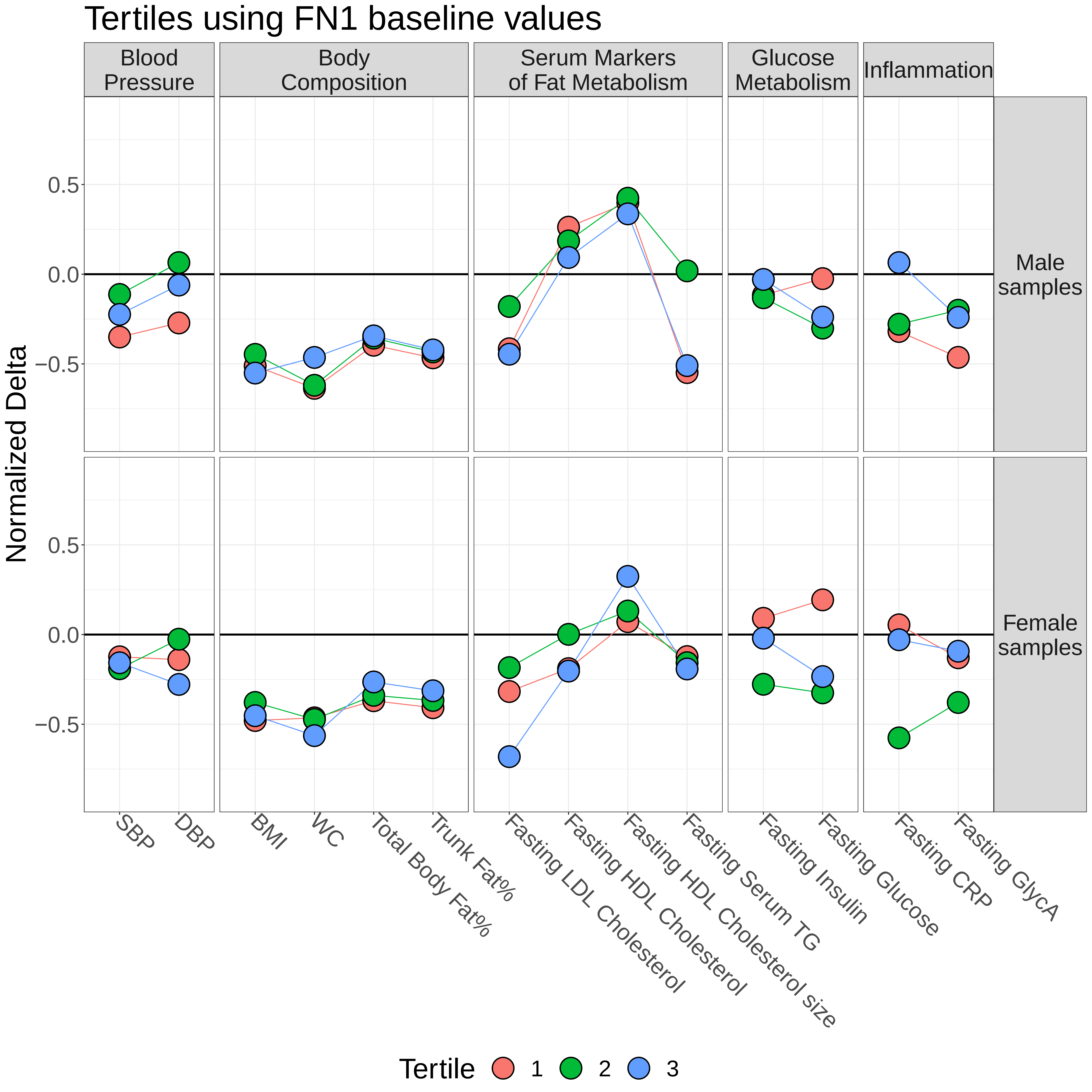

### Supplementary Figure 13

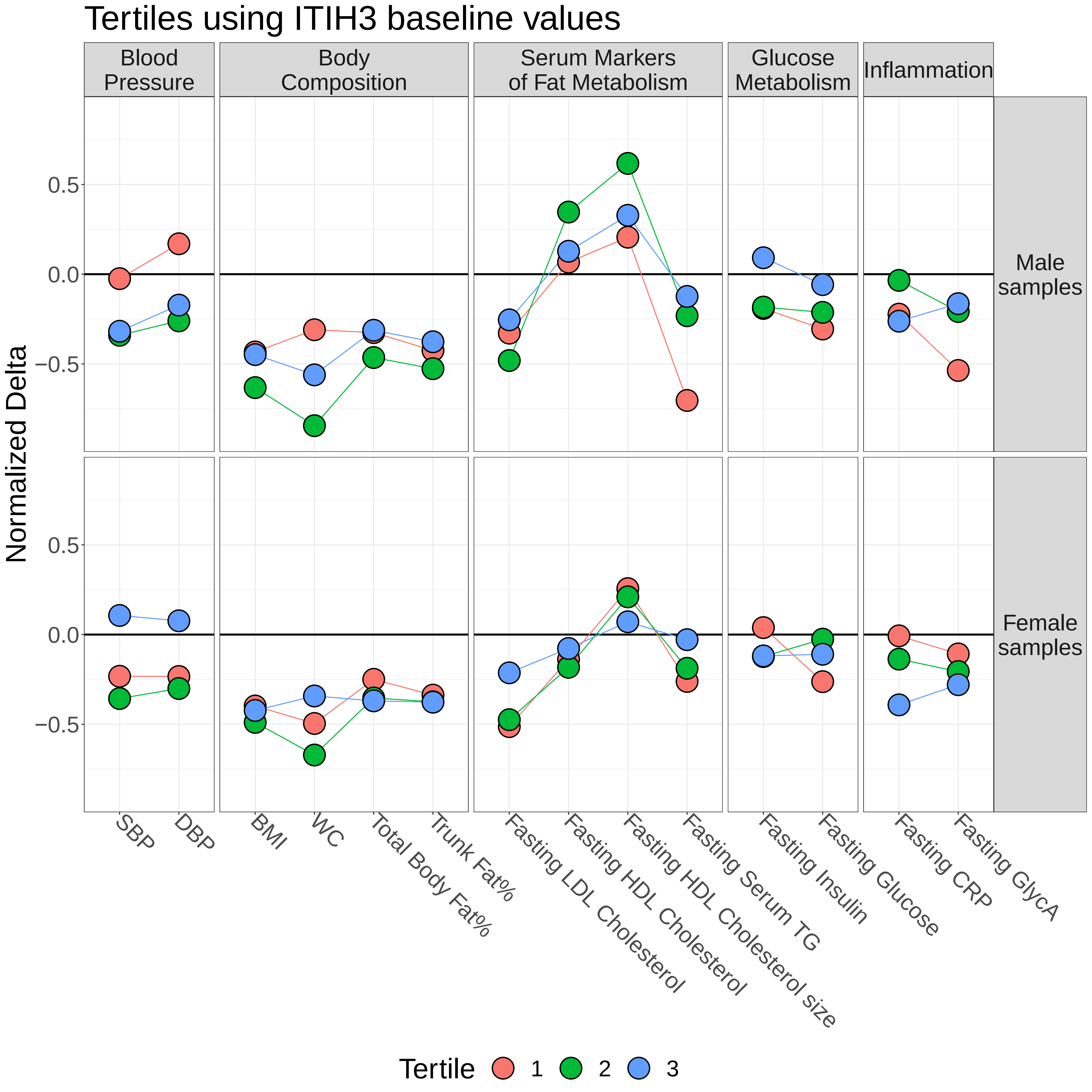

### Supplementary Figure 14

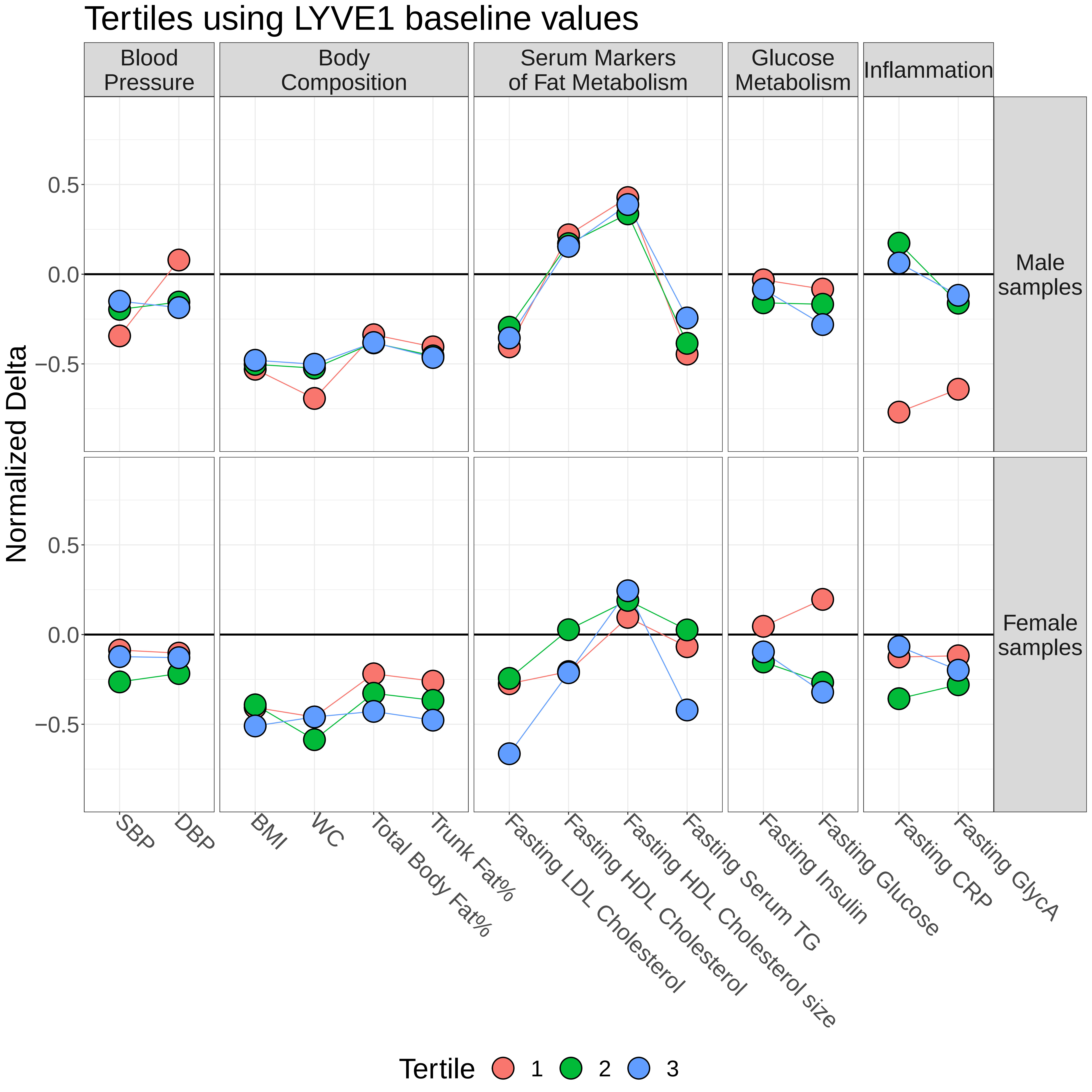

### Supplementary Figure 15

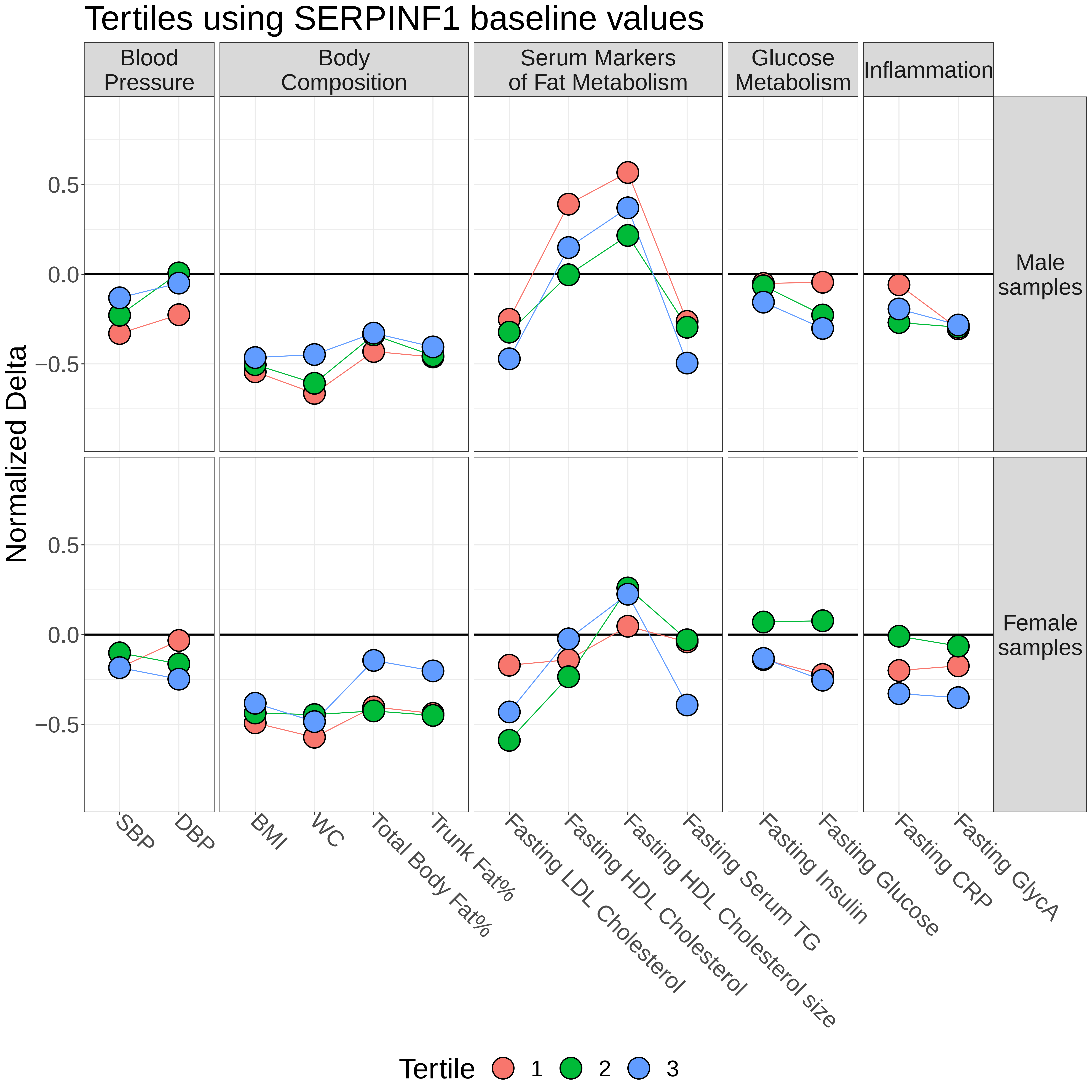

### Supplementary Figure 16

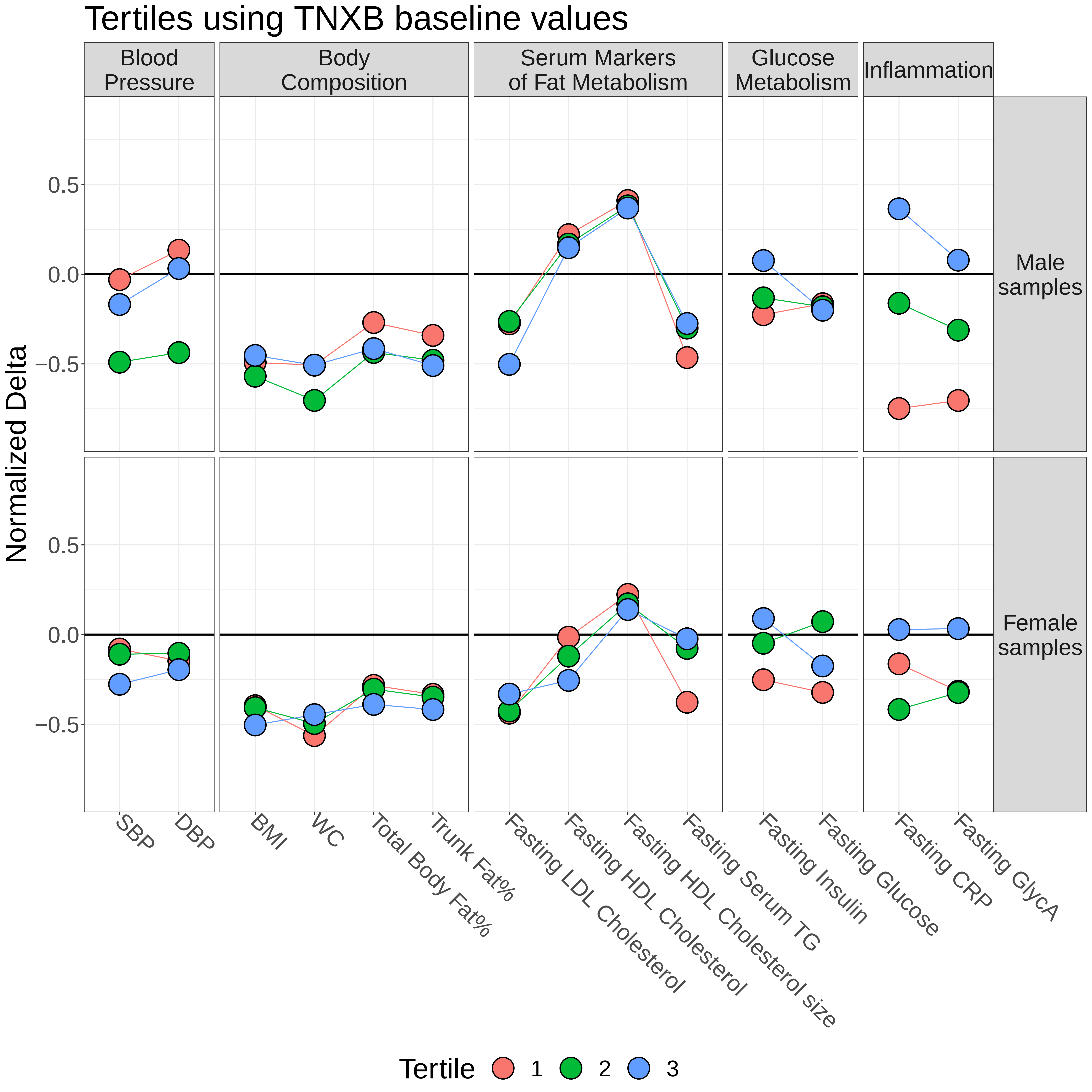
